# Computational Exploration of Phytochemicals as Potent Inhibitors of Acetylcholinesterase Enzyme in Alzheimer’s Disease

**DOI:** 10.1101/2020.01.04.20016535

**Authors:** Bishajit Sarkar, Md. Asad Ullah, Md. Nazmul Islam Prottoy

## Abstract

Alzheimer’s Disease (AD) is the most common type of age related dementia in the world. Many hypotheses shed light on several reasons that lead to AD development. The cholinergic hypothesis describes that the destruction of an essential neurotransmitter, acetylcholine (AChE) by acetylcholinesterase (AChE) enzyme, leads to the AD onset. The hydrolysis of acetylcholine by excess amount of AChE decreases the amount of acetylcholine in the brain, thus interfering with the normal brain functions. Many anti-AChE agents can be used to treat AD by targeting AChE. In our study, 14 anti-AChE agents from plants: 1,8-cineol, berberine, carvacrol, cheilanthifoline, coptisine, estragole, harmaline, harmine, liriodenine, myrtenal, naringenin, protopine, scoulerine, stylopine were tested against AChE and compared with two controls: donepezil and galantamine, using different techniques of molecular docking. Molecular docking study was conducted for all the 14 selected ligands against AChE to identify the best three ligands among them. To determine the safety and efficacy of the three best ligands, a set of tests like the druglikeness property test, ADME/T test, PASS & P450 site of metabolism prediction, pharmacophore mapping and modelling and DFT calculations were performed. In our experiment, berberine, coptisine and naringenin were determined as the three ligands from the docking study. Further analysis of these 3 ligands showed coptisine as the most potent anti-AChE agent. The molecular dynamics simulation study showed quite good stability of the coptisine-AChE docked complex. Administration of berberine, coptisine and naringenin might be potential treatments for AD.

## 1. Introduction

Alzheimer’s Disease (AD) was first described by Alois Alzheimer in 1907. It is one of the most prevalent dementia type diseases as well as a common type of age related dementia that is increasing its numbers day by day ^[1, 2]^. The common symptoms of AD include intellectual morbidity, delusions, psychomotor dysregulation, hallucinations etc. ^[3]^. Genetic factors play key roles in the familial cases of AD ^[4]^. There are many reasons that lead to the AD development. Many hypotheses have been developed by the scientists that indicate several reasons for the AD onset. One hypothesis is called the amyloid cascade hypothesis, where the deposition of β-amyloid plaques in the brain is responsible for the development of AD. Abnormal processing of amyloid precursor protein (APP) by β-secretase enzyme produces the β-amyloid plaques in the brain. Studies have showed that these plaques interfere with the normal brain functions ^[5]^. Moreover, another hypothesis, called the oxidative stress hypothesis, describes that because of the deposition of increased amount of iron, aluminium and mercury, free radicals are generated very rapidly and events like lipid peroxidation, protein and DNA oxidation increase dramatically in the brain. The stresses produced by these oxidation events are responsible for the development of AD ^[6]^. According to another hypothesis called cholinergic hypothesis, the loss of the functions of cholinergic neurons and cholinergic neurotransmission in the brain is responsible for AD ^[7]^. Our study was conducted focusing on the cholinergic hypothesis of AD development.

### 1.1. The cholinergic hypothesis and AD development

The cholinergic hypothesis involves one of the major neurotransmitters, acetylcholine and its regulation by two enzymes, acetylcholinesterase and choline acetyltransferase ^[8]^. Acetylcholine (ACh) is a major neurotransmitter that mediates many important functions of the brain including the learning and memory processes. ACh mediates its effects through binding to two types of receptors i.e., nicotinic (α7 and α4β2) and muscarinic receptors (M1 muscarinic receptor). The AChE is synthesized by an enzyme called choline-acetyltransferase (ChAT). ChAT catalyzes the transfer of the acetyl group from acetyl coenzyme A (AcCoA) to choline (Ch) in the pre-synaptic neuron and thus synthesize the ACh. The ACh is then secreted by the pre-synaptic neuron into the synapse, which then mediates its effects by binding to either the nicotinic receptor or muscarinic receptor. To maintain the proper concentration of ACh in the brain, an enzyme called acetylcholinesterase (AChE), is synthesized. This enzyme is a serine hydrolase that hydrolyzes ACh to acetate and Ch. The Ch is again taken up by the pre-synaptic neuron for recycling and reusing. In this way, the balance of ACh is maintained in the normal brain. However, there is evidence that, in the brain of AD patients, the overexpression of AChE occurs. Due to this reason, the break-down of AChE occurs rapidly, which decreases the required amount of ACh in the brain. Due to the scarcity of enough ACh in the brain, the neuron cells cannot function properly and brain damage as well as memory loss occur, which lead to the onset of AD development (**Figure 01**). The AChE inhibitors repress or inhibit the activity of AChE. For this reason, AChE can be a potential target for anti-AChE drugs to treat AD (**Figure 01**) ^[9, 10, 11, 12, 13]^.

**Figure 01.**
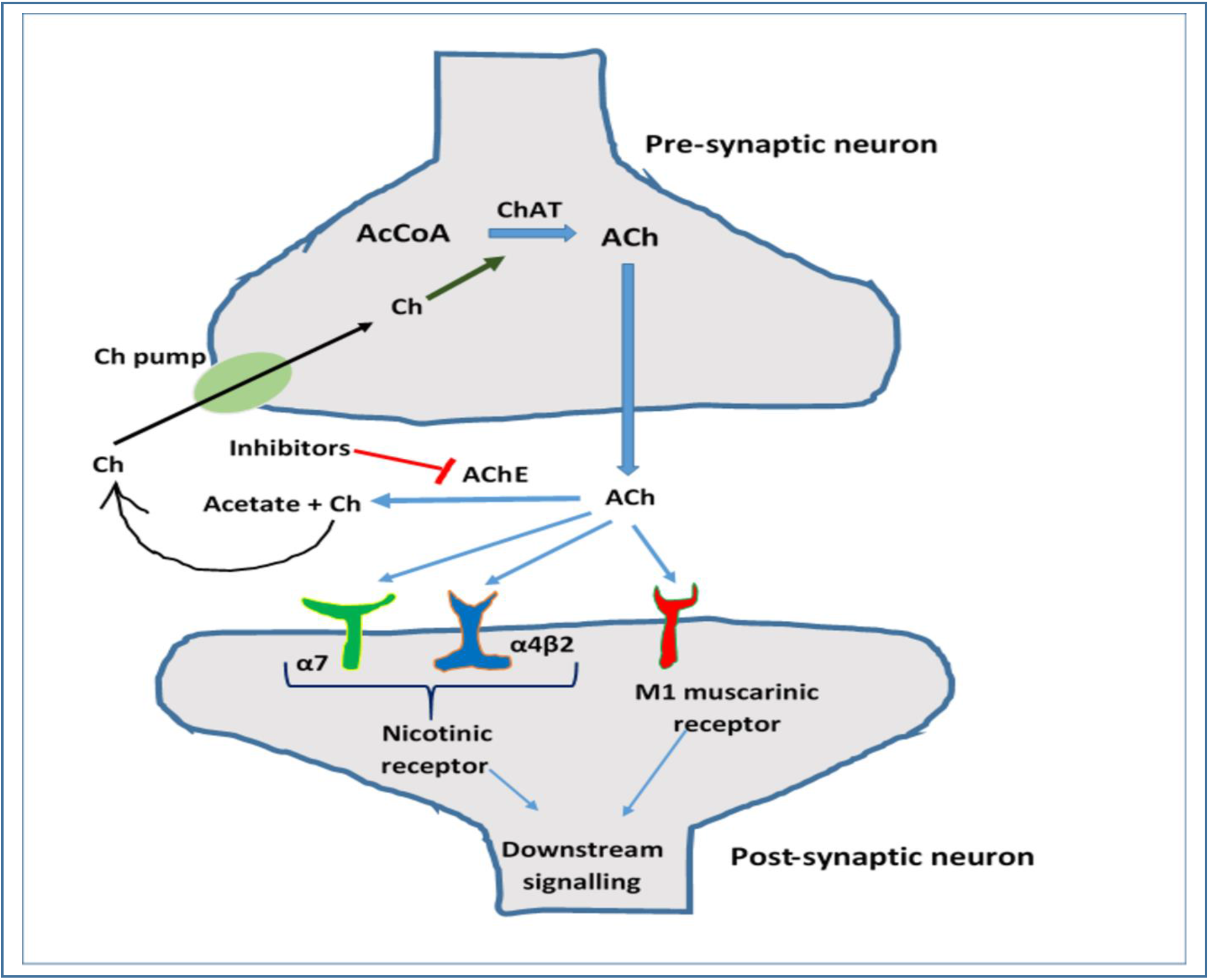
Cholinergic hypothesis and role of acetylcholinesterase in AD development. Acteylcholine (ACh) is synthesized (synthesizing reaction is catalysed by choline acetyltransferase, ChAT) and released by the pre-synaptic neuron. The AChE mediates its effects on the post-synaptic neuron through nicotinic and/or muscarinic receptors. The ACh later performs the downstream signalling in the post-synaptic neuron. AChE enzyme breaks down the ACh to acetate and choline (Ch) and so the overexpression of AChE lowers the amount of ACh in the brain which leads to the AD onset. AChE inhibitors repress the AChE activity, thus aid in the AD treatment.

### 1.2. Anti-acetylcholinesterase agents from plants

Plants have a long history to be used in different types of medical purposes ^[14, 15]^. Recently, various anti-AChE agents have been identified in the plants. Many of these natural agents show good efficacy in inhibiting the AChE activity ^[16, 17]^. In our study, 14 potential anti-AChE agents were selected to analyze their inhibitory activities against the AChE enzyme as well as their safety and efficacy, using the techniques of molecular docking. The 14 agents are listed in **Table 01** along with their source plants. Molecular docking, also known as computational drug design, is a widely accepted and used technique for new lead discovery. This technique reduces both time and costs of the drug discovery processes. Till now, over 50 drugs have been designed with the aid of computational simulation tools and many of them have received FDA approval for marketing. Molecular docking tries to predict the pose, interaction and conformation of a ligand molecule within the binding pocket of a target molecule by mimicking or simulating the actual biological environment in the computer software. After estimating the type of interactions, the software assigns scoring function to each of the bound ligands that reflects their binding affinity. The lower scores corresponds to greater binding affinity. These scores are predicted by specific algorithms of the software ^[18, 19]^. Along with the molecular docking study, ADME/T tests are also performed to identify the safety and efficacy of a candidate drug molecule ^[20]^. The main objective of this study is to compare and identify three best agents from the group of 14 anti-AChE agents. At first, the molecular docking study was conducted for all the 14 selected ligands against the AChE enzyme (PDB ID: 1ACJ). Based on the docking scores, three best molecules were selected. Later, druglikeness property experiments, ADME/T tests, PASS prediction, P450 site of metabolism prediction, pharmacophore mapping and modelling and DFT calculations were performed on the three best selected ligands using various tools to determine their safety and efficacy. A flowchart of the steps of the study is depicted in **Figure 02**. Molecular docking study using different ligands has already been performed against the AChE enzyme (PDB ID: 1ACJ), where satisfied results were obtained ^[21]^. Two FDA approved drugs i.e., donepezil and galantamine, were used as positive controls in this study. These two drugs are also AChE inhibitors. Galantamine is approved for treating the mild and moderate AD and dopenezil is used to treat mild, moderate and severe AD ^[22]^.

**Table 01.**
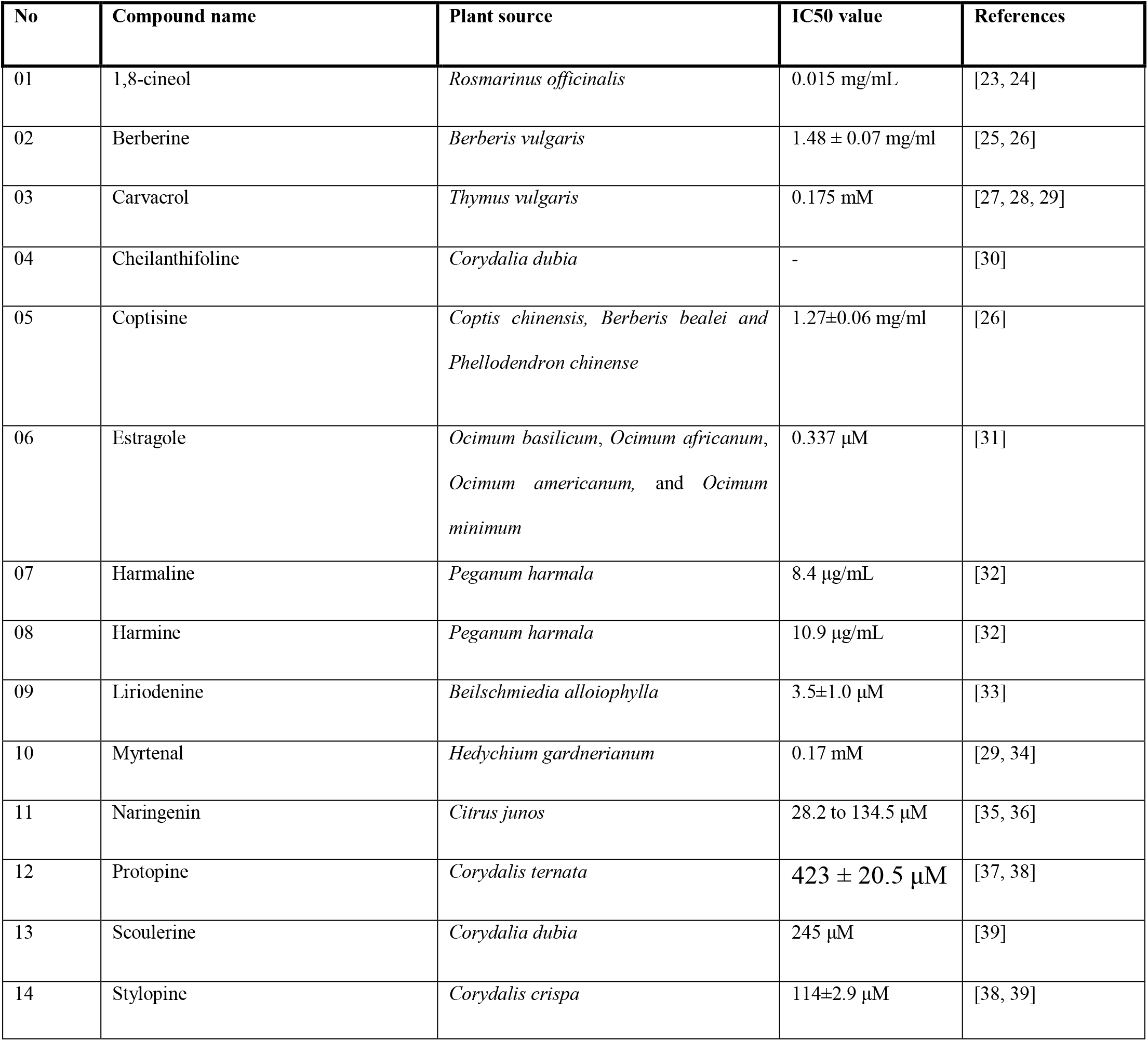
List of the 14 anti-AChE agents from plants used in the study, with their IC50 values.

**Figure 02.**
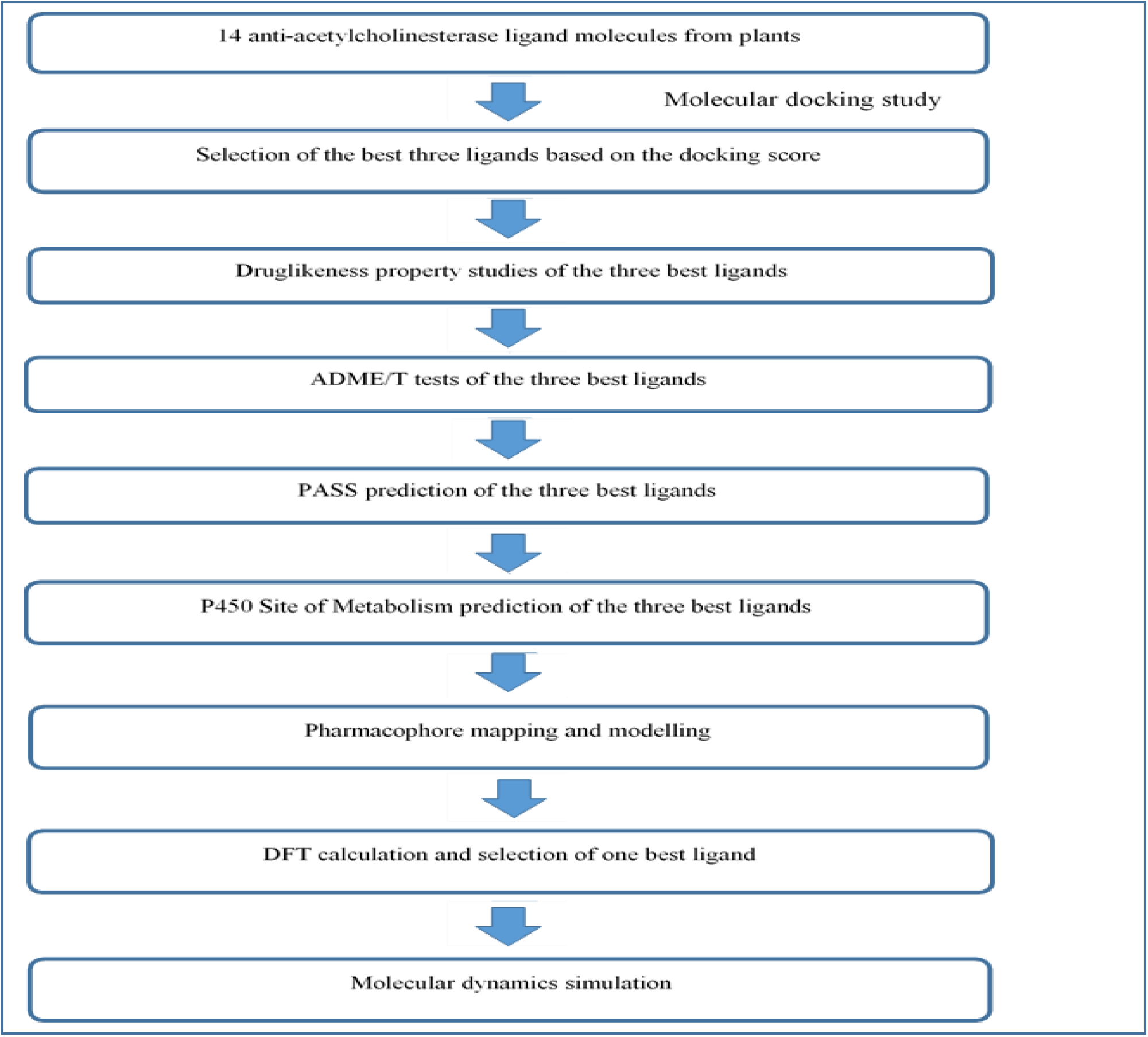
The flowchart of the work-plan of the experiment.

## 2. Materials and Methods

Ligand preparation, grid generation and glide docking between the ligands and their receptor, AChE were obtained using Maestro-Schrödinger Suite 2018-4. The 2D and 3D representations of the best pose interactions between the best three ligands and AChE were visualized using Maestro-Schrödinger Suite 2018-4 and Discovery Studio Visualize, respectively ^[40, 41]^.

### 2.1. Protein Preparation

Three dimensional structure of AChE enzyme (PDB ID: 1ACJ) was downloaded in PDB format from protein data bank (www.rcsb.org) **(Figure 03)**. The proteins were then prepared and refined using the Protein Preparation Wizard in Maestro Schrödinger Suite 2018-4 ^[42]^. All the water molecules were deleted from the protein during protein preparation. Finally, the structure was optimized and then minimized using force field OPLS_2005. Minimization was done setting the maximum heavy atom RMSD (root-mean-square-deviation) to 30 Å and any remaining water less than 3 H-bonds to non-water was again deleted during the minimization step.

**Figure 03.**
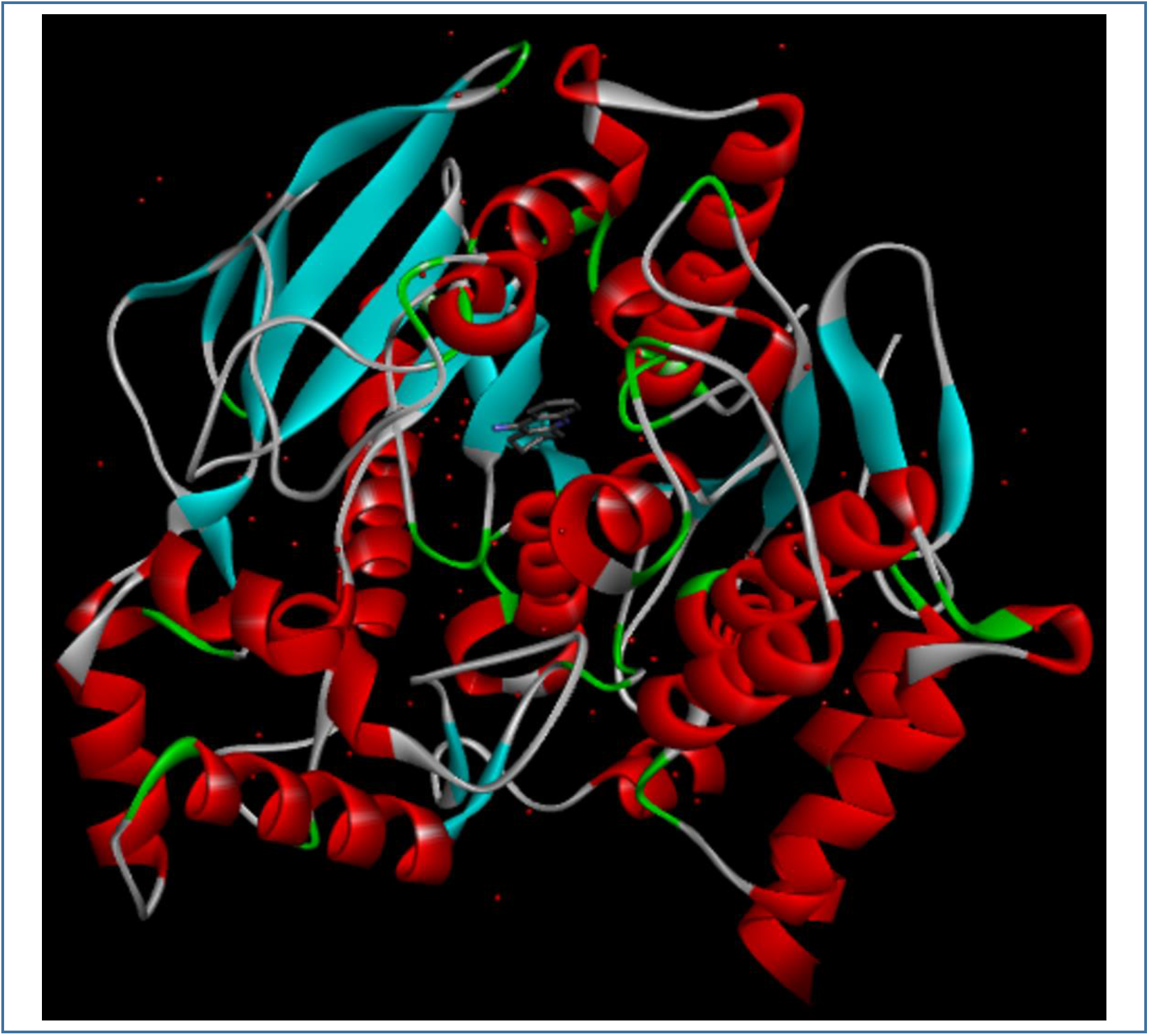
3D structure of AChE enzyme (PDB ID: 1ACJ). The structure was taken from Protein Data Bank and visualized using Discovery Studio Visualizer.

### 2.2. Ligand Preparation

Structures of the controls: donepezil (PubChem CID: 3152) and galantamine (PubChem CID: 9651) and the selected ligands: 1,8-cineol (PubChem CID: 2758), berberine (PubChem CID: 2353), carvacrol (PubChem CID: 10364), cheilanthifoline (PubChem CID: 440582), coptisine (PubChem CID: 72322), estragole (PubChem CID: 8815), harmaline (PubChem CID: 3564), harmine (PubChem CID: 5280953), liriodenine (PubChem CID: 10144), myrtenal (PubChem CID: 61130), naringenin (PubChem CID: 932), protopine (PubChem CID: 4970), scoulerine (PubChem CID: 22955), stylopine (PubChem CID: 6770) were downloaded in SDF format (sequentially) from PubChem (www.pubchem.ncbi.nlm.nih.gov) **(Figure 04)**. These structures were then prepared using the LigPrep function of Maestro Schrödinger Suite 2018-4 ^[43]^. Minimized 3D structures of ligands were generated using Epik2.2 and within pH 7.0 +/- 2.0. Minimization was again carried out using OPLS_2005 force field which generated 32 possible stereoisomers.

**Figure 04.**
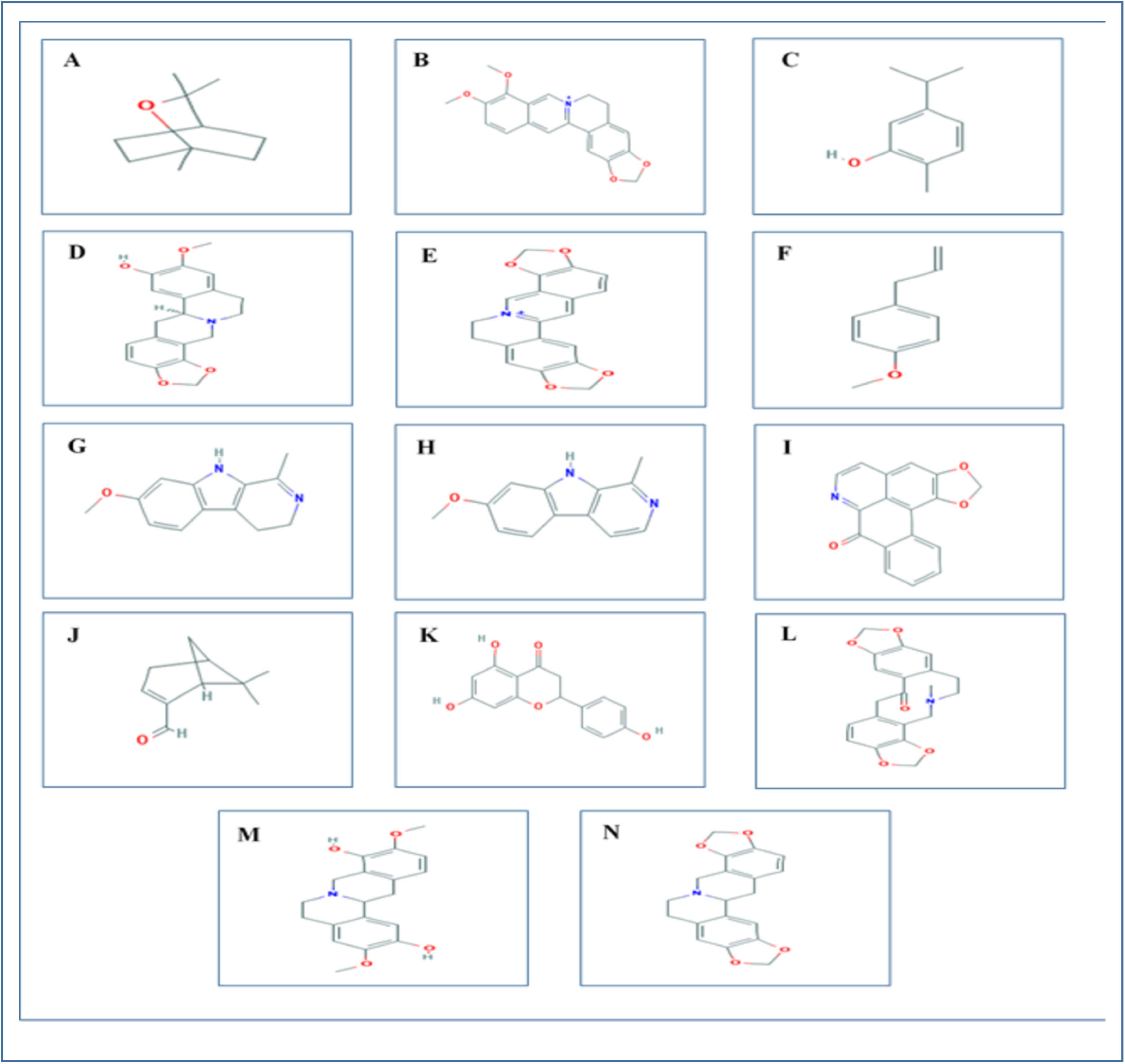
2D representations of the all the 14 ligands used in the experiment. The ligand structures were taken from PubChem server (www.pubchem.ncbi.nlm.nih.gov. A. 1,8-cineol; B. Berberine; C. Carvacrol; D. Cheilanthifoline; E. Coptisine; F. Estragole; G. Harmaline; H. Harmine; I. Liriodenine; J. Myrtenal; K. Naringenin; L. Protopine; M. Scoulerine; N. Stylopine.

### 2.3. Receptor Grid Generation

Grid usually confines the active site to shortened specific area of the receptor protein for the ligand to dock specifically. In Glide, a grid was generated using default Van der Waals radius scaling factor 1.0 and charge cutoff 0.25 which was then subjected to OPLS_2005 force field. A cubic box was generated around the active site (reference ligand active site). Then the grid box volume was adjusted to 15×15×15 for docking test.

### 2.4. Glide Standard Precision (SP) and Extra Precision (XP) Ligand Docking, Prime MM-GBSA Prediction and Induced Fit Docking

SP and XP glide docking were carried out using Glide in Maestro Schrödinger Suite 2018-4 ^[44]^. The Van der Waals radius scaling factor and charge cutoff were set to 0.80 and 0.15 respectively for all the ligand molecules. Final score was assigned according to the pose of docked ligand within the active site of the receptor. The ligand with the lowest glide docking score was considered as the best ligand. The docking results are listed in **Table 02**. After successful docking, the 2D representations of the best pose interactions between the three best ligands and their receptor were generated using Maestro-Schrödinger Suite 2018-4 **(Figure 05)**. The 3D representations of the best pose interactions between the three best ligands and their receptor were obtained using Discovery Studio Visualizer **(Figure 06)**. The interaction of the best three ligands with different amino acids of the receptor protein was also visualized by the Discovery Studio Visualizer **(Figure 07)**. The molecular mechanics-generalized born and surface area (MM-GBSA) tool was used to determine the ΔG_Bind_ scores and induced fit docking (IFD) was carried out to predict the XP G_Score_ scores of only the three best ligand molecules. Both the MM-GBSA and IFD studies were carried out using Maestro-Schrödinger Suite 2018-4. To determine the ΔG_Bind_ scores of the best three ligands, OPLS_2005 was selected as the force field as well as the VSGB solvation model was selected. The other parameters were kept default. To carry out the IFD study, the protein preparation was first carried out and then the OPLS_2005 was selected as the force field, rigid docking was selected in the conformational sampling parameter, receptor van der Waals screening was kept 0.70, ligand van der Waals screening was set at 0.50 and maximum number of poses was set at 2, refine residues within 2 angstrom of ligand poses and Extra Precision (XP) were selected. Other parameters were kept default. The results of MM-GBSA (ΔG_Bind_ scores) study and IFD (XP G_Score_ and IFD values) study are listed in **Table 03**. Moreover, **Table 03** also lists the different types of bonds and bond distances that took part in the interaction of the three best ligands and their receptor, AChE. After the docking analysis, three best ligand molecules were chosen and then they were further analyzed for various physicochemical properties.

**Figure 05.**
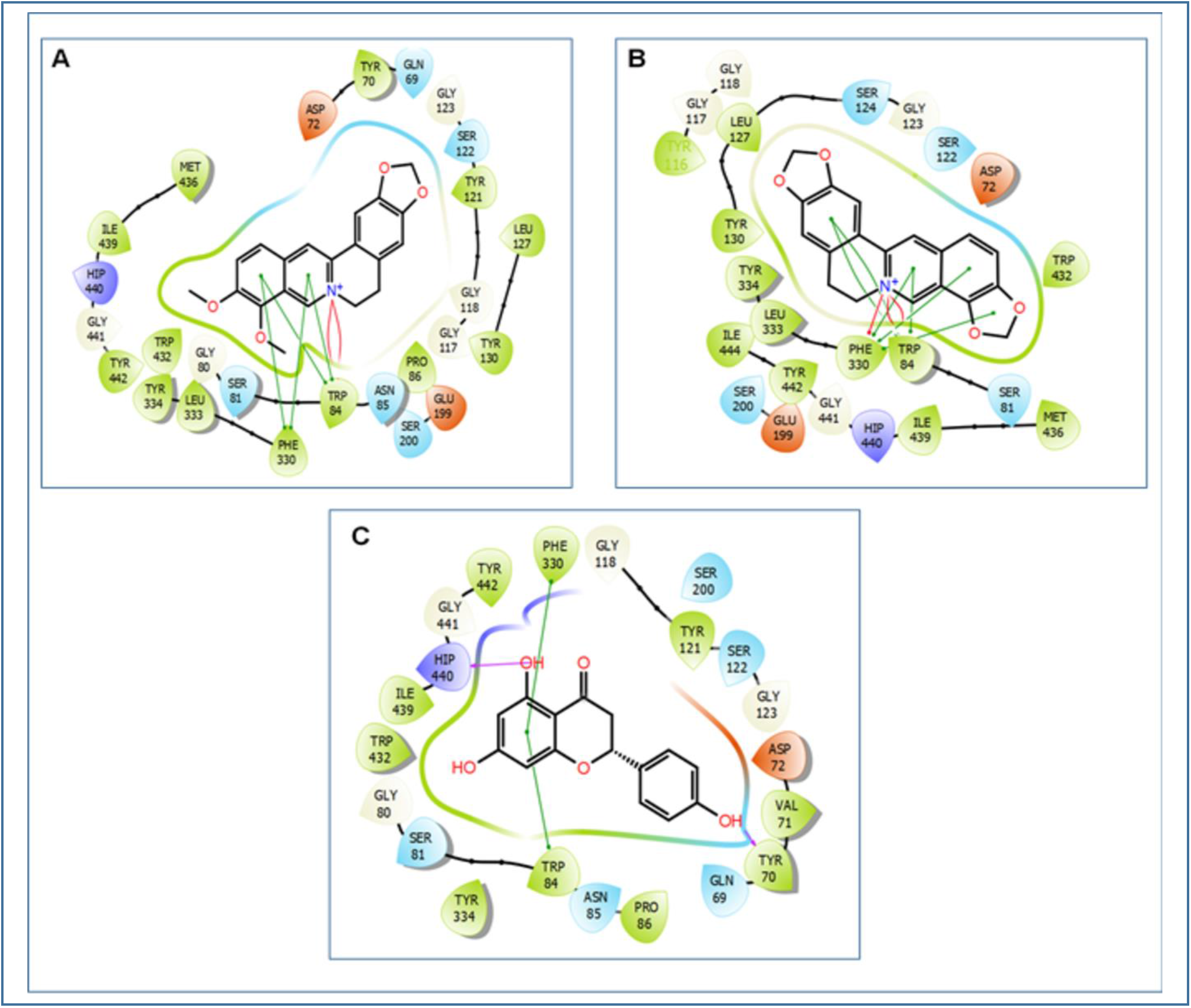
2D representations of the best pose interactions between the three best ligands and their receptor. A. interaction between berberine and AChE, B. interaction between coptisine and AChE, C. interaction between naringenin and AChE. Colored spheres indicates the type of residue in the target: red-negatively charged (Asp, Glu), blue-polar (Ser, Gln, Asn), green-hydrophobic (Tyr, Met, Leu, Trp, Ile, Phe, Pro), ash color-glycine, deep purple-unspecified molecules and the greyish circles represent solvent exposure. Interactions are shown as colored lines-solid pink lines with arrow-H-bond in target (backbone), dotted pink lines with arrow-H-bond between receptor and ligand (sidechain), solid pink lines without arrow-metal co-ordination, green line-pi-pi stacking interaction, green dotted lines-distances, partially blue and red colored lines- salt bridges. Ligands exposed to solvent are represented by grey spheres. The colored lines show the protein pocket for the ligand according to nearest atom. Interruptions of the lines indicate the opening of the pocket. 2D representations of the best pose interactions between the ligands and their respective receptors were obtained using Maestro-Schrödinger Suite 2018-4.

**Figure 06.**
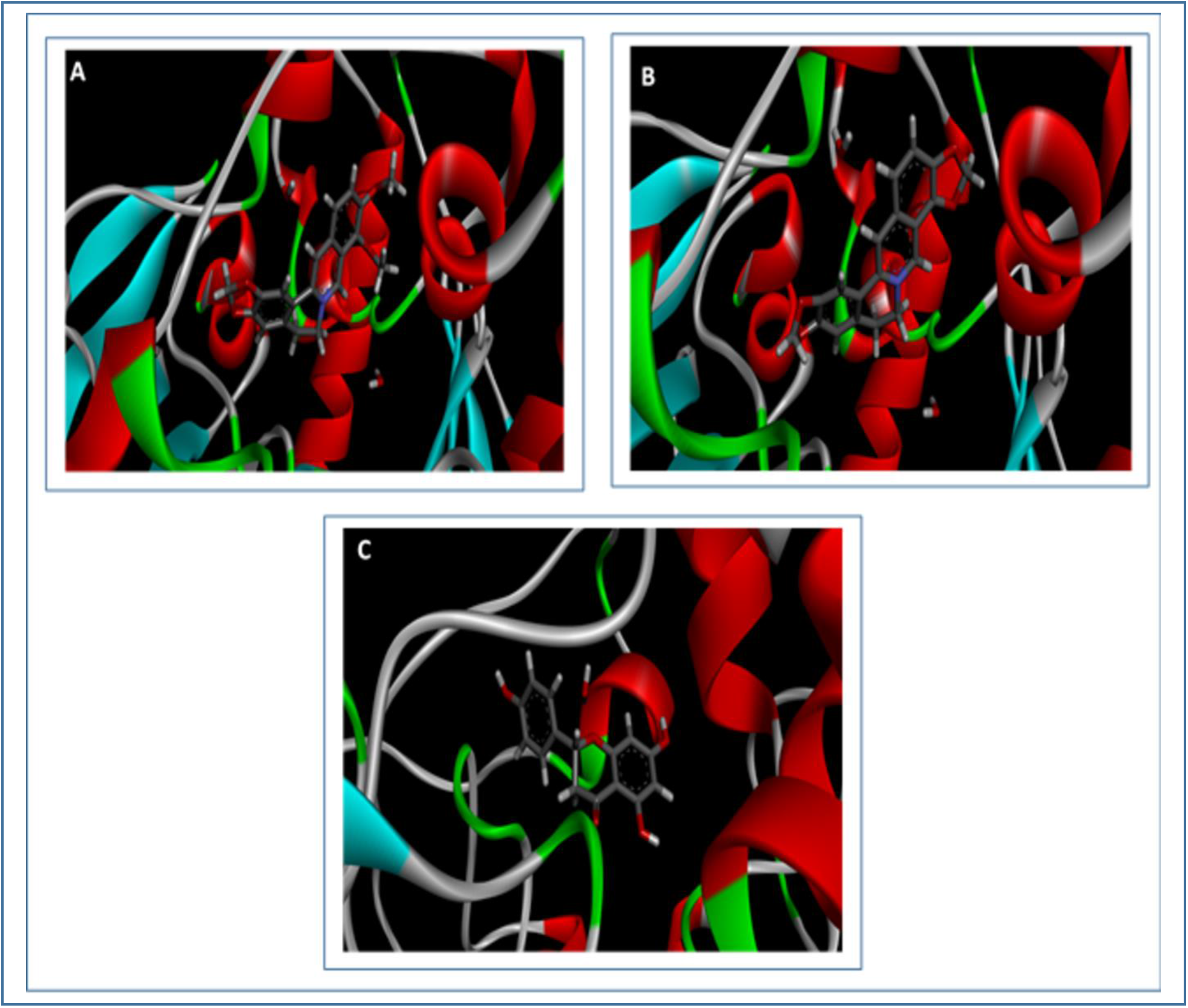
3D representations of the best pose interactions between the ligands and their receptor. The proteins are represented in solid ribbon model and the ligands are represented in stick model. A. interaction between berberine and AChE, B. interaction between coptisine and AChE, C. interaction between naringenin and AChE. The 3D representations of the best pose interactions between the ligands and their respective receptors were visualized using Discovery Studio Visualizer.

**Figure 07.**
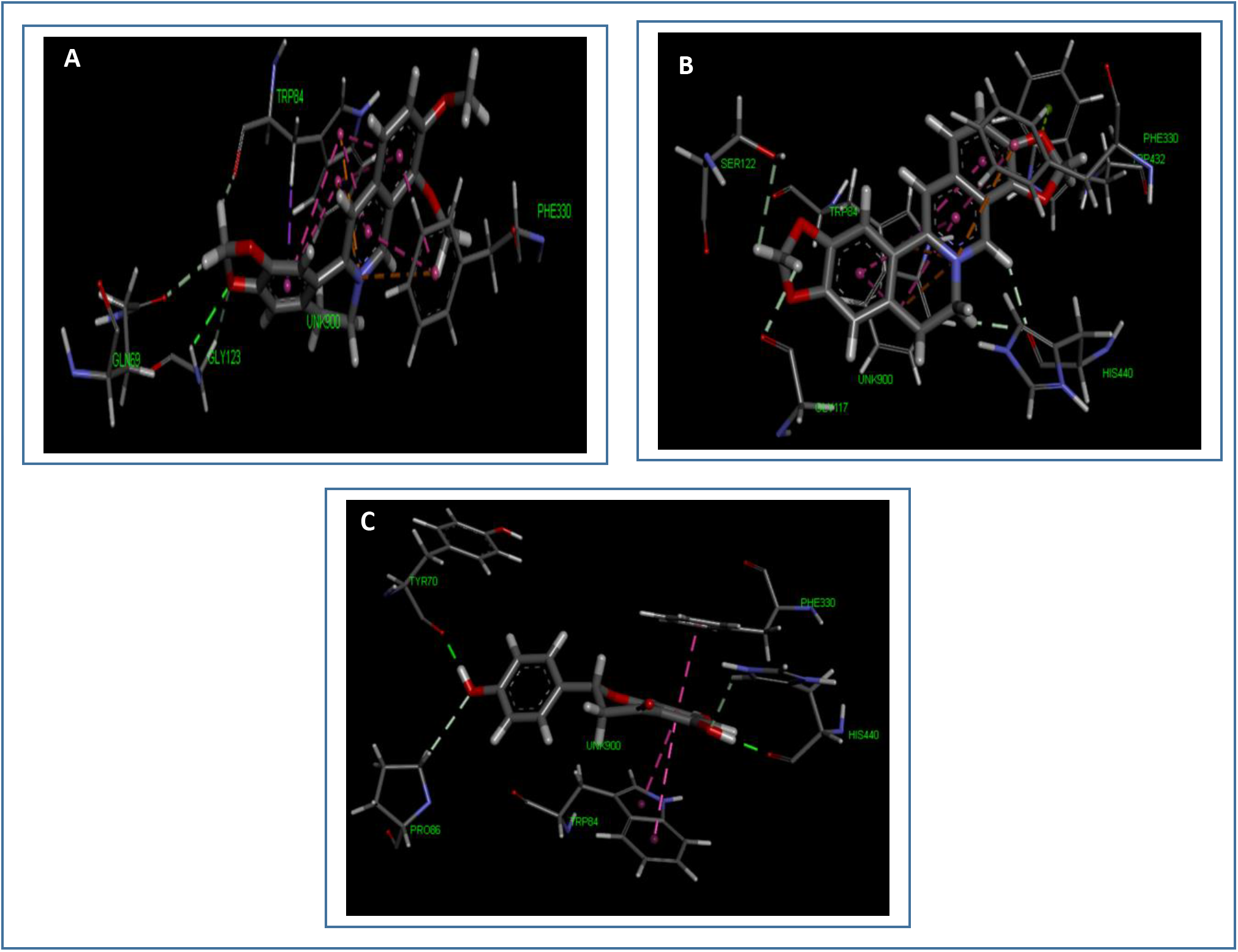
Figure showing the various types of bonds and amino acids that take part in the interaction between the three best selected ligands and their receptor. Interacting amino acid residues of target molecule are labeled in the diagram and dotted lines depict interaction between ligand and receptor. Green dotted lines- conventional bond, light pink- alkyl/pi-alkyl interactions, yellow- pi-sulfur/sulphur-X interaction, deep pink- pi-pi stacked bond, orange- charge-charge interaction, purple- pi-sigma interaction, red- donor-donor interaction. A. interaction between berberine and AChE, B. interaction between coptisine and AChE, C. interaction between naringenin and AChE.

**Table 02.** The docking results (binding energy) of all the 14 ligands and the controls along with the determination of Lipinski’s rule of five, their respective number of hydrogen bonds as well as interacting amino acids.

| No | Names of ligands<br>(with PubChem<br>CID) | SP Docking Score<br>(Binding Energy)<br>(Kcal/mol) | XP Docking<br>Score (Binding<br>Energy)<br>(Kcal/mol) | Lipinski's rule<br>of five | Interacting residues of<br>the target |
| --- | --- | --- | --- | --- | --- |
| 01 | 1,8-cineol<br>(PubChem CID:<br>2758) | -4.844 | -3.109 | Yes | Trp 84, Phe 330 |
| 02 | Berberine<br>(PubChem CID:<br>2353) | -9.658 | -13.571 | Yes | Gln 69, Phe 330, Gly 123,<br>Trp 84 |
| 03 | Carvacrol<br>(PubChem CID:<br>10364) | -6.060 | -6.986 | Yes | Tyr 334, Phe 330, Trp 432,<br>Tyr 442, Trp 84, His 440,<br>Arg 289 |
| 04 | Cheilanthifoline<br>(PubChem CID:<br>440582) | -6.387 | -7.398 | Yes | Gly 117, Trp 84, Phe 330,<br>Trp 432 |
| 05 | Coptisine<br>(PubChem CID:<br>72322) | -10.148 | -15.560 | Yes | Trp 432, Trp 84, Phe 330,<br>His 440, Ser 122, Gly 117 |
| 06 | Estragole<br>(PubChem CID:<br>8815) | -5.035 | -5.992 | Yes | Tyr 334, Phe 330, Ile 439,<br>Trp 432, Trp 84 |
| 07 | Harmaline<br>(PubChem CID:<br>3564) | -8.053 | -9.154 | Yes | Tyr 334, Trp 432, Trp 84,<br>Glu 199, Tyr 442, His 440,<br>Ile 439, Phe 330 |
| 08 | Harmine (PubChem<br>CID: 5280953) | -8.385 | -8.363 | Yes | Phe 330, His 440, Trp 84 |
| 09 | Liriodenine<br>(PubChem CID:<br>10144) | -7.754 | -7.775 | Yes | Phe 330, Tyr 334, Tyr 121,<br>Trp 84 |
| 10 | Myrtenal (PubChem<br>CID: 61130) | -5.873 | -4.821 | Yes | Ile 444, Ser 200, Gly 118,<br>Gly 119, Trp 84, Phe 330,<br>His 440 |
| 11 | Naringenin<br>(PubChem CID:<br>932) | -9.266 | -9.342 | Yes | His 440, Phe 330, Tyr 70,<br>Trp 84, Pro 86 |
| 12 | Protopine<br>(PubChem CID:<br>4970) | -6.272 | -7.789 | Yes | Phe 330, His 440, Gly 117,<br>Trp 84 |
| 13 | Scoulerine<br>(PubChem CID:<br>22955) | -6.229 | -2.230 | Yes | Phe 330, His 440, Tyr 130,<br>Gly 117, Trp 84, Ser 122 |
| 14 | Stylopine<br>(PubChem CID:<br>6770) | -7.733 | -8.071 | Yes | Phe 330, His 440, Trp 84,<br>Gly 117 |
| Control 1 | Donepezil<br>(PubChem CID:<br>3152) | -5.045 | -8.434 | Yes | Trp 279, Tyr 70, Arg 289 |
| Control 2 | Galantamine<br>(PubChem CID:<br>9651) | -7.516 | -7.237 | Yes | Trp 84, Glu 199, His 440,<br>Phe 330, Tyr 121, Tyr 70,<br>Gly 117 |

**Table 03.** List of the different types of bonds along with their bond distances with their respective amino acids, that took part in the interaction between the three best ligands as well as the controls and the target receptor, AChE, along with their ΔG_Bind_ scores, XP G_Score_, IFD scores and glide energy.

| Name of the ligand (with respective receptor) | MM-GBSA ($\Delta G_{\text{Bind}}$ Score Kcal/mol) | XP $G_{\text{Score}}$ (Kcal/mol) | IFD score (Kcal/mol) | Glide energy (Kcal/mol) | Interacting amino acids | Bond distance in Å | Interaction category | Type of interaction |
| --- | --- | --- | --- | --- | --- | --- | --- | --- |
| Berberine | -35.80 | -12.889 | -1154.880 | -23.549 | Glutamine 69 | 2.22 | Hydrogen | Carbon |
|  |  |  |  |  | Glycine 123 | 2.82 | Hydrogen | Carbon |
|  |  |  |  |  |  | 2.42 | Hydrogen | Conventional |
|  |  |  |  |  | Tryptophan 84 | 2.22 | Hydrogen | Carbon |
|  |  |  |  |  |  | 2.79 | Hydrophobic | Pi-Sigma |
|  |  |  |  |  |  | 3.95 | Hydrophobic | Pi-Pi stacked |
|  |  |  |  |  |  | 5.43 | Hydrophobic | Pi-Pi stacked |
|  |  |  |  |  |  | 3.68 | Hydrophobic | Pi-Pi stacked |
|  |  |  |  |  |  | 4.62 | Hydrophobic | Pi-Pi stacked |
|  |  |  |  |  |  | 5.92 | Hydrophobic | Pi-Pi stacked |
|  |  |  |  |  |  | 5.42 | Hydrophobic | Pi-Pi stacked |
|  |  |  |  |  |  | 4.41 | Electrostatic | Pi-Cation |
|  |  |  |  |  |  | 4.12 | Electrostatic | Pi-Cation |
|  |  |  |  |  | Phenylalanine 330 | 3.85 | Hydrophobic | Pi-Pi stacked |
|  |  |  |  |  |  | 4.02 | Hydrophobic | Pi-Pi stacked |
|  |  |  |  |  |  | 4.64 | Electrostatic | Pi-Cation |
| Coptisine | -49.91 | 14.942 | -1158.410 | -16.197 | Tryptophan 432 | 2.96 | Hydrophobic | Pi-Lone pair |
|  |  |  |  |  | Tryptophan 84 | 4.48 | Hydrophobic | Pi-Pi stacked |
|  |  |  |  |  |  | 3.62 | Hydrophobic | Pi-Pi stacked |
|  |  |  |  |  |  | 4.53 | Hydrophobic | Pi-Pi stacked |
|  |  |  |  |  |  | 4.33 | Hydrophobic | Pi-Pi stacked |
|  |  |  |  |  |  | 4.02 | Hydrophobic | Pi-Pi stacked |
|  |  |  |  |  |  | 3.94 | Electrostatic | Pi-Cation |
|  |  |  |  |  |  | 4.12 | Electrostatic | Pi-Cation |
|  |  |  |  |  | Phenylalanine 330 | 3.84 | Hydrophobic | Pi-Pi stacked |
|  |  |  |  |  |  | 3.74 | Hydrophobic | Pi-Pi stacked |
|  |  |  |  |  |  | 4.43 | Electrostatic | Pi-Cation |
|  |  |  |  |  | Histidine 440 | 2.21 | Hydrogen | Carbon |
|  |  |  |  |  |  | 2.13 | Hydrogen | Carbon |
|  |  |  |  |  | Serine 122 | 3.01 | Hydrogen | Carbon |
|  |  |  |  |  | Glycine 117 | 2.53 | Hydrogen | Carbon |
| Naringenin<br>(PubChem<br>CID: 932) | -23.73 | -10.078 | -1152.980 | -45.657 | Histidine 440 | 1.69 | Hydrogen | Conventional |
|  |  |  |  |  |  | 2.38 | Hydrogen | Carbon |
|  |  |  |  |  | Phenylalanine<br>330 | 3.60 | Hydrophobic | Pi-Pi stacked |
|  |  |  |  |  | Tyrosine 70 | 1.66 | Hydrogen | Conventional |
|  |  |  |  |  | Tryptophan 84 | 3.84 | Hydrophobic | Pi-Pi stacked |
|  |  |  |  |  |  | 4.78 | Hydrophobic | Pi-Pi stacked |
|  |  |  |  |  | Proline 86 | 2.99 | Hydrogen | Carbon |
| Donepezil<br>(PubChem<br>CID: 3152) | -47.06 | -8.209 | -1148.970 | -33.756 | Tryptophan 279 | 5.23 | Hydrophobic | Pi-Alkyl |
|  |  |  |  |  | Tyrosine 70 | 5.32 | Hydrophobic | Pi-Pi stacked |
|  |  |  |  |  |  | 2.25 | Hydrogen | Carbon |
|  |  |  |  |  | Arginine 289 | 2.21 | Hydrogen | Carbon |
| Galantamine<br>(PubChem<br>CID: 9651) | -5.19 | -5.748 | -1147.050 | -33.736 | Tryptophan 84 | 4.78 | Hydrophobic | Pi-Alkyl |
|  |  |  |  |  |  | 4.07 | Hydrophobic | Pi-Alkyl |
|  |  |  |  |  |  | 5.37 | Hydrophobic | Pi-Alkyl |
|  |  |  |  |  |  | 2.21 | Hydrophobic | Pi-Pi sigma |
|  |  |  |  |  |  | 2.56 | Hydrogen | Carbon |
|  |  |  |  |  | Glutamic acid<br>199 | 2.57 | Hydrogen | Carbon |
|  |  |  |  |  |  | 2.49 | Hydrogen | Carbon |
|  |  |  |  |  | Histidine 440 | 1.96 | Hydrogen | Conventional |
|  |  |  |  |  | Phenylalanine<br>330 | 4.30 | Hydrophobic | Pi-Alkyl |
|  |  |  |  |  | Tyrosine 121 | 2.42 | Hydrogen | Carbon |
|  |  |  |  |  | Tyrosine 70 | 2.87 | Hydrogen | Carbon |
|  |  |  |  |  | Glycine 117 | 4.19 | Hydrophobic | Amide-Pi stacked |

### 2.5. Ligand Based Drug Likeness Property and ADME/Toxicity Prediction

The molecular structures of the three best ligands were analyzed using SWISSADME server (http://www.swissadme.ch/). In the druglikeness property test, Lipinki’s rule of five or not, along with some other properties were predicted. Various physicochemical properties of ligand molecules were calculated using OSIRIS Property Explorer. The drug likeness properties of the selected ligand molecules were analyzed using SWISSADME server as well as the OSIRIS Property Explorer ^[45, 46]^. The results of drug likeness property analysis are summarized in **Table** 04^[47]^. The ADME/T study for each of the ligand molecules was carried out using an online based server ADMETlab (http://admet.scbdd.com/) to predict their various pharmacokinetic and pharmacodynamic properties including blood brain barrier permeability, human intestinal adsorption, Caco-2 permeability, Cytochrome P (CYP) inhibitory capability, half-life, mutagenicity etc. ^[48]^. The numeric and categorical values of the results generated by ADMETlab tool were changed into qualitative values according to the explanation and interpretation described in the ADMETlab server (http://admet.scbdd.com/home/interpretation/) for the convenience of interpretation. The results of ADME/T for all the ligand molecules are depicted in **Table 05**.

**Table 04.** List of the results of the druglikeness properties of the three best ligands: berberine, coptisine and naringenin and the controls.

| Drug Likeness Properties | Berberine | Coptisine | Naringenin | Donepezil<br>(control 1) | Galantamine<br>(control 2) |
| --- | --- | --- | --- | --- | --- |
| Molecular weight | 336.36 g/mol | 320.32 g/mol | 272.25 g/mol | 379.49 g/mol | 287.35 g/mol |
| Consensus Log $P_{o/w}$ | 2.53 | 2.40 | 1.84 | 4.00 | 1.91 |
| Log $S$ | -4.55 | -4.52 | -3.49 | -4.81 | -2.93 |
| Num. H-bond acceptors | 4 | 4 | 5 | 4 | 4 |
| Num. H-bond donors | 0 | 0 | 3 | 0 | 1 |
| Molar Refractivity | 94.87 | 87.95 | 71.57 | 115.31 | 84.05 |
| Lipinski | Yes | Yes | Yes | Yes | Yes |
| Ghose | Yes | Yes | Yes | Yes | Yes |
| Veber | Yes | Yes | Yes | Yes | Yes |
| Egan | Yes | Yes | Yes | Yes | Yes |
| Muegge | Yes | Yes | Yes | Yes | Yes |
| Bioavailability score | 0.55 | 0.55 | 0.55 | 0.55 | 0.55 |
| Synthetic accessibility (SA) | 3.14 | 2.96 | 3.01 | 3.62 | 4.57 |
| TPSA ($\text{\AA}^2$) | 40.80 | 40.80 | 71.57 | 38.77 | 41.93 |
| No of rotatable bonds | 2 | 0 | 1 | 6 | 1 |
| Druglikeness score | - | - | 1.9 | 7.29 | 6.2 |
| Drug-Score | - | - | 0.84 | 0.63 | 0.91 |
| Solubility | - | - | -2.64 | -4.35 | -2.67 |
| Reproductive effective | - | - | No | No | No |
| Irritant | - | - | No | No | No |
| Tumorigenic | - | - | No | No | No |
| Mutagenic | - | - | No | No | No |

**Table 05.** The ADME/T test results of the best three ligand molecules and the controls. The tests were carried out using ADMETlab server.

| Class | Properties | Berberine | Coptisine | Naringenin | Donepezil<br>(control 1) | Galantamine<br>(control 2) |
| --- | --- | --- | --- | --- | --- | --- |
| Absorption | Caco-2 permeability | Optimal | Optimal | Optimal | Optimal | Optimal |
|  | Pgp-inhibitor | Non-inhibitor | Non-inhibitor | Non-inhibitor | Inhibitor | Non-inhibitor |
|  | Pgp-substrate | Substrate | Non-substrate | Non-substrate | Substrate | Substrate |
|  | Human Intestinal Absorption<br>(HIA) | HIA negative | HIA negative | HIA positive | HIA<br>positive | HIA positive |
| Distribution | Plasma Protein Binding | Low | Low | Good | Optimal | Low |
|  | BBB (Blood–Brain Barrier) | BBB positive | BBB positive | BBB positive | BBB<br>positive | BBB positive |
| Metabolism | CYP450 1A2 inhibitor | Inhibitor | Inhibitor | Inhibitor | Non-inhibitor | Non-inhibitor |
|  | CYP450 1A2 substrate | Substrate | Substrate | Substrate | Non-substrate | Substrate |
|  | CYP450 3A4 inhibitor | Inhibitor | Non-inhibitor | Inhibitor | Non-inhibitor | Non-inhibitor |
|  | CYP450 3A4 substrate | Substrate | Substrate | Non-substrate | Substrate | Substrate |
|  | CYP450 2C9 inhibitor | Non-inhibitor | Non-inhibitor | Non-inhibitor | Non-inhibitor | Non-inhibitor |
|  | CYP450 2C9 substrate | Non-substrate | Non-substrate | Non-substrate | Non-substrate | Non-substrate |
|  | CYP450 2C19 inhibitor | Non-inhibitor | Non-inhibitor | Non-inhibitor | Non-inhibitor | Non-inhibitor |
|  | CYP450 2C19 substrate | Substrate | Substrate | Non-substrate | Substrate | Non-substrate |
|  | CYP450 2D6 inhibitor | Inhibitor | Inhibitor | Non-inhibitor | Inhibitor | Inhibitor |
|  | CYP450 2D6 substrate | Substrate | Substrate | Non-substrate | Substrate | Substrate |
| Excretion | T<sub>1/2</sub> (h) | 1.9 | 1.8 | 0.9 | 1.7 | 1.7 |
| Toxicity | hERG (hERG Blockers) | Blocker | Blocker | Blocker | Blocker | Non-blocker |
|  | H-HT (Human Hepatotoxicity) | HHT positive | HHT negative | HHT negative | HHT positive | HHT positive |
|  | Ames (Ames Mutagenicity) | Ames negative | Ames negative | Ames negative | Ames negative | Ames negative |
|  | DILI (Drug Induced Liver Injury) | DILI positive | DILI positive | DILI positive | DILI negative | DILI negative |

### 2.6. PASS (Prediction of Activity Spectra for Substances) and P450 Site of Metabolism (SOM) prediction

The PASS (Prediction of Activity Spectra for Substances) prediction of the three best selected ligands were conducted using PASS-Way2Drug server (http://www.pharmaexpert.ru/passonline/) by using canonical SMILES from PubChem server (https://pubchem.ncbi.nlm.nih.gov/) ^[49]^. To carry out PASS prediction, P_a_ (probability “to be active”) was kept greater than 70% because studies have confirmed that the P_a_ > 70% threshold gives highly reliable prediction ^[50]^. In the PASS prediction study, both possible biological activities and possible adverse and toxic effects of the selected ligands were predicted. **Table 06** and **Table 07** list the results of the PASS prediction studies. The P450 site of metabolism (SOM) of the three best selected ligand molecules were determined by online tool, RS-WebPredictor 1.0 (http://reccr.chem.rpi.edu/Software/RS-WebPredictor/) ^[51]^. The LD50 values of the best three ligands were determined from admetSAR (http://lmmd.ecust.edu.cn/admetsar2/) server. The server predicts the acute oral toxicity class from the canonical SMILES of a compound and the LD50 value can be derived from the predicted category ^[52]^. The category-I represents the compounds with LD50 values less than or equal to 50mg/kg, category-II represents the compounds with LD50 values greater than 50mg/kg but less than 500mg/kg, Category-III contains compounds with LD50 values greater than 500mg/kg but less than 5000mg/kg and category-IV is comprised of the compounds with LD50 values greater than 5000mg/kg ^[53]^. The toxicity classes of the best three compounds were predicted using ProTox-II server (http://tox.charite.de/protox_II/) ^[54]^. The canonical SMILES of berberine, coptisine and naringenin were taken from PubChem server (https://pubchem.ncbi.nlm.nih.gov/) and the SMILES were used to predict the toxicity class. **Table 08** lists the results of P450 SOM study.

**Table 06.** The results of the PASS prediction study of the biological activities of the best three ligand molecules.

| Sl no | Biological activities | Berberine |  | Coptisine |  | Naringenin |  |
| --- | --- | --- | --- | --- | --- | --- | --- |
|  |  | Predicted LD50: More than 500 mg/kg |  | Predicted LD50: More than 500 mg/kg |  | Predicted LD50: More than 50 mg/kg |  |
|  |  | Toxicity class: 4 |  | Toxicity class: 4 |  | Toxicity class: 4 |  |
|  |  | Pa | Pi | Pa | Pi | Pa | Pi |
| 01 | Membrane integrity agonist | - | - | - | - | 0.964 | 0.003 |
| 02 | HMOX1 expression enhancer | - | - | - | - | 0.956 | 0.002 |
| 03 | Chlordecone reductase inhibitor | - | - | - | - | 0.918 | 0.004 |
| 04 | HIF1A expression inhibitor | - | - | - | - | 0.911 | 0.005 |
| 05 | Histidine kinase inhibitor | - | - | - | - | 0.892 | 0.002 |
| 06 | Aldehyde oxidase inhibitor | - | - | - | - | 0.868 | 0.005 |
| 07 | Antimutagenic | - | - | - | - | 0.857 | 0.003 |
| 08 | Mucomembranous protector | - | - | - | - | 0.844 | 0.010 |
| 09 | TP53 expression enhancer | - | - | - | - | 0.822 | 0.009 |
| 10 | Chemopreventive | - | - | - | - | 0.724 | 0.006 |

**Table 07.**
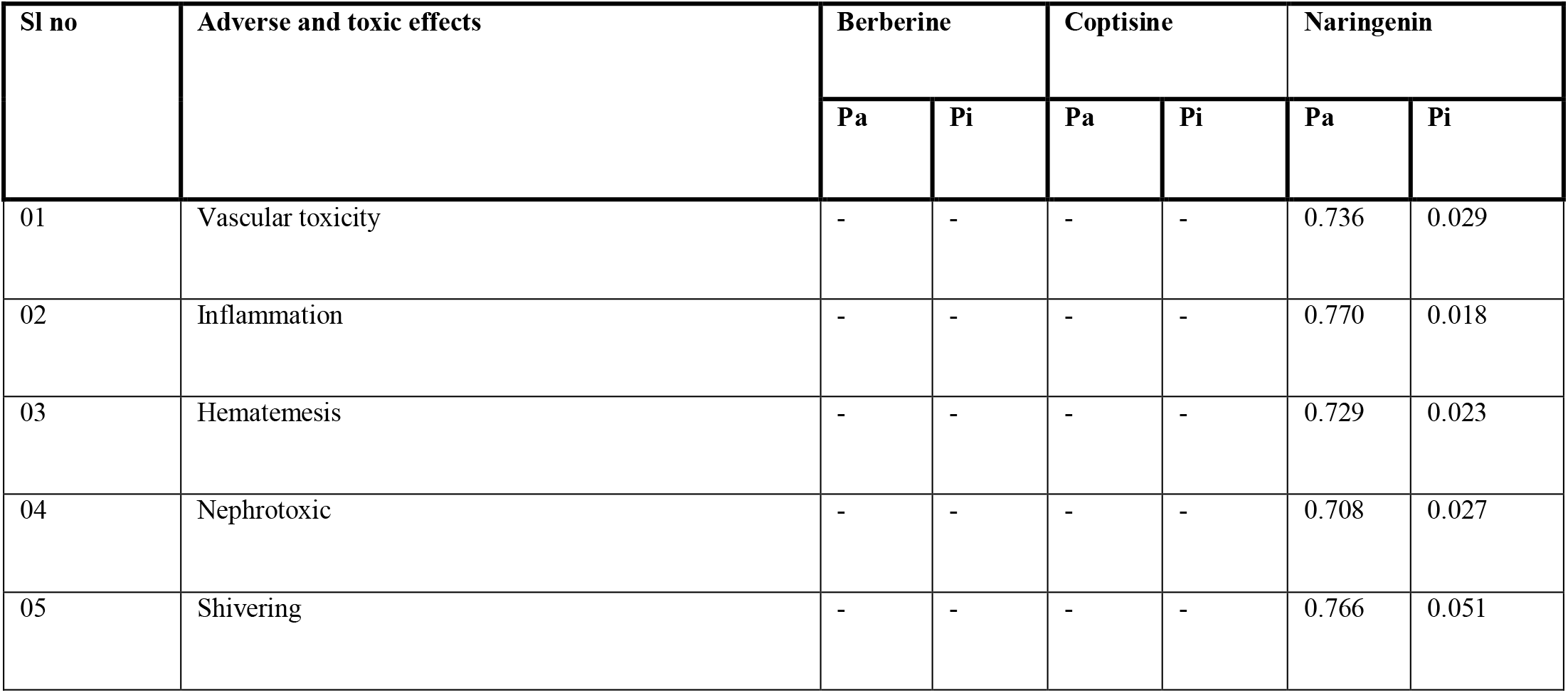
The results of the PASS prediction study showing the adverse and toxic effects of the best three ligand molecules.

**Table 08.**
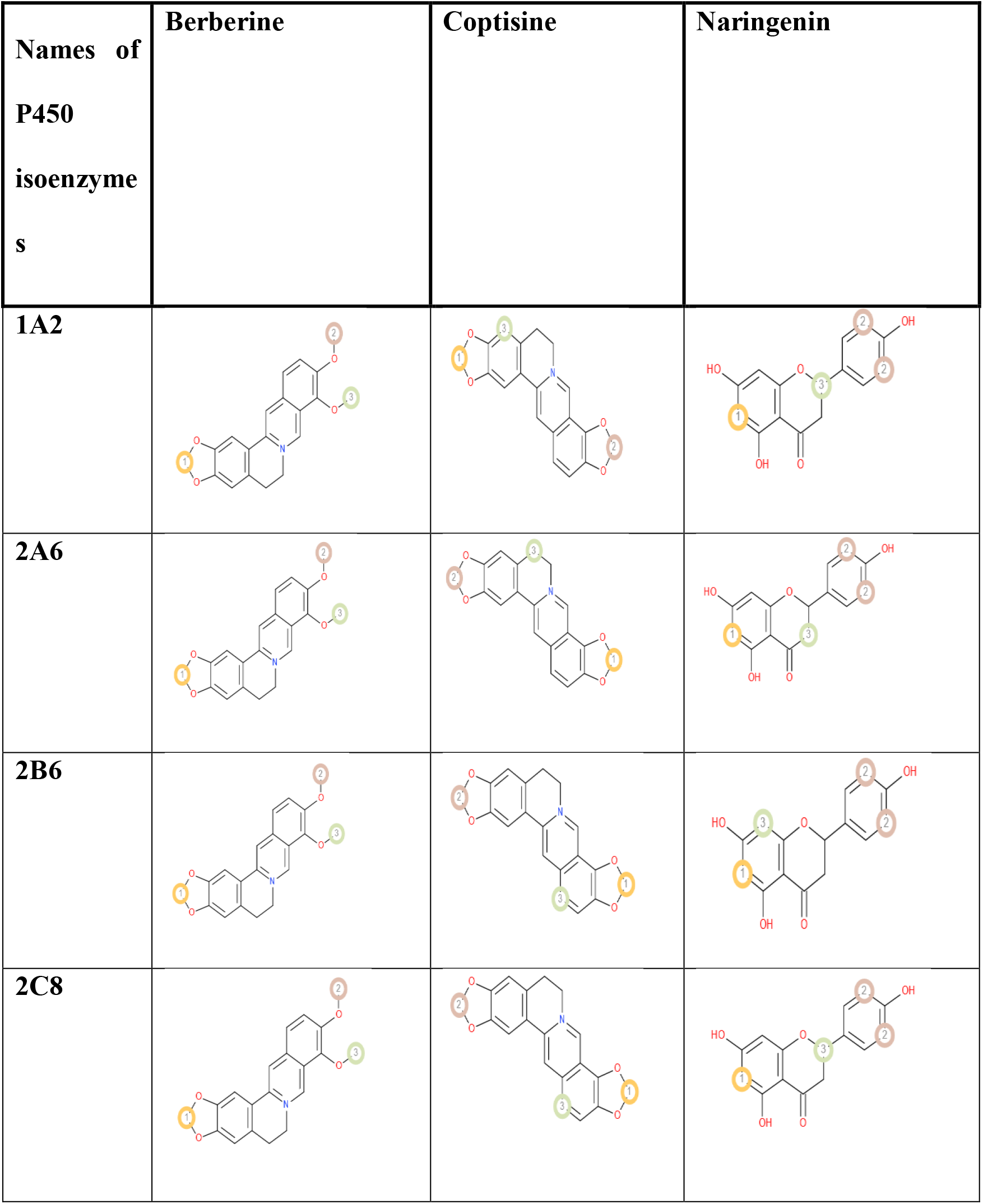
Results of the P450 sites of metabolism prediction study of the three best ligand molecules.

**Table 09.**
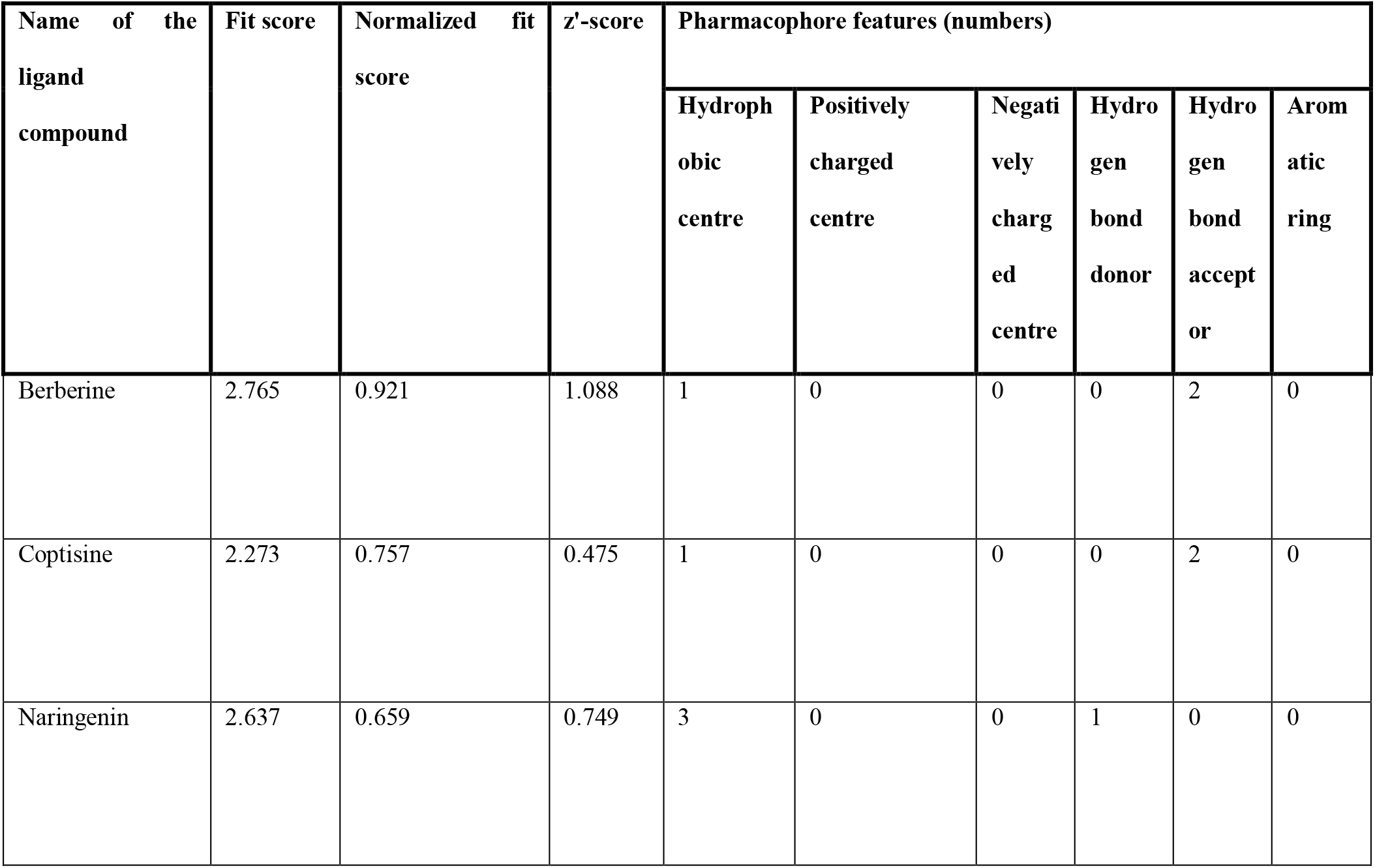
Results of the pharmacophore mapping experiment of the three best ligands, berberine, coptisine and naringenin.

### 2.7. Pharmacophore Mapping and Modelling

The pharmacophore mapping study of the three best ligands was carried out by online server PharmMapper (http://www.lilab-ecust.cn/pharmmapper/) ^[55]^. The ligands, downloaded in SDF format from PubChem server, were uploaded and the “maximum number of conformations” parameter was set at 1000, all possible targets were kept at the “select target set” parameter and the “number of reserved matched targets” parameter was kept 1000. In the advanced options, the cut-off value of fit score was set at 0. All the other parameters were kept default. The pharmacophore mapping experiment was done for the three best ligand molecules among the 14 selected ligands (**Figure 08 and Table 09**)

**Figure 08.**
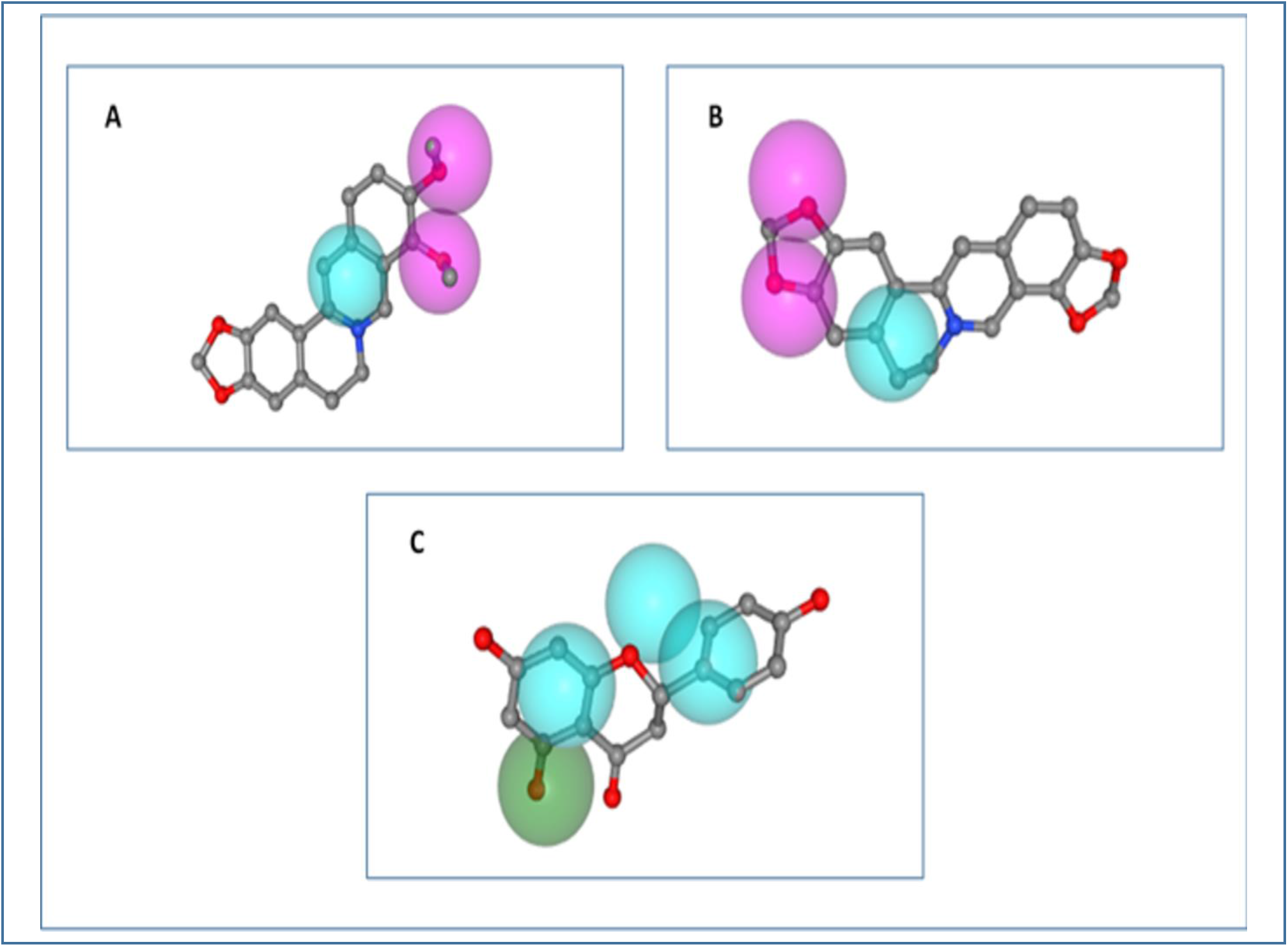
Parmacophore mapping of A. berberine, B. coptisine, C. naringenin. Here, light blue colour- hydrophobic centre, green colour- hydrogen bond donor and pink colour- hydrogen bond acceptor.

The pharmacophore modelling of the three best ligands was performed using the Phase pharmacophore perception engine of Maestro-Schrödinger Suite 2018-4 ^[56]^. The pharmacophore modelling was done manually, where the radii sizes were kept as the Van der Waals radii of receptor atoms, radii scaling factor was kept at 0.50, receptor atoms whose surfaces were within 2.00 Å of the ligand surface were ignored and the volume shell thickness was limited to 5.00 Å. The 2D and 3D pharmacophore modelling were carried out for the three best ligand molecules (**Figure 09 and Figure 10**).

**Figure 09.**
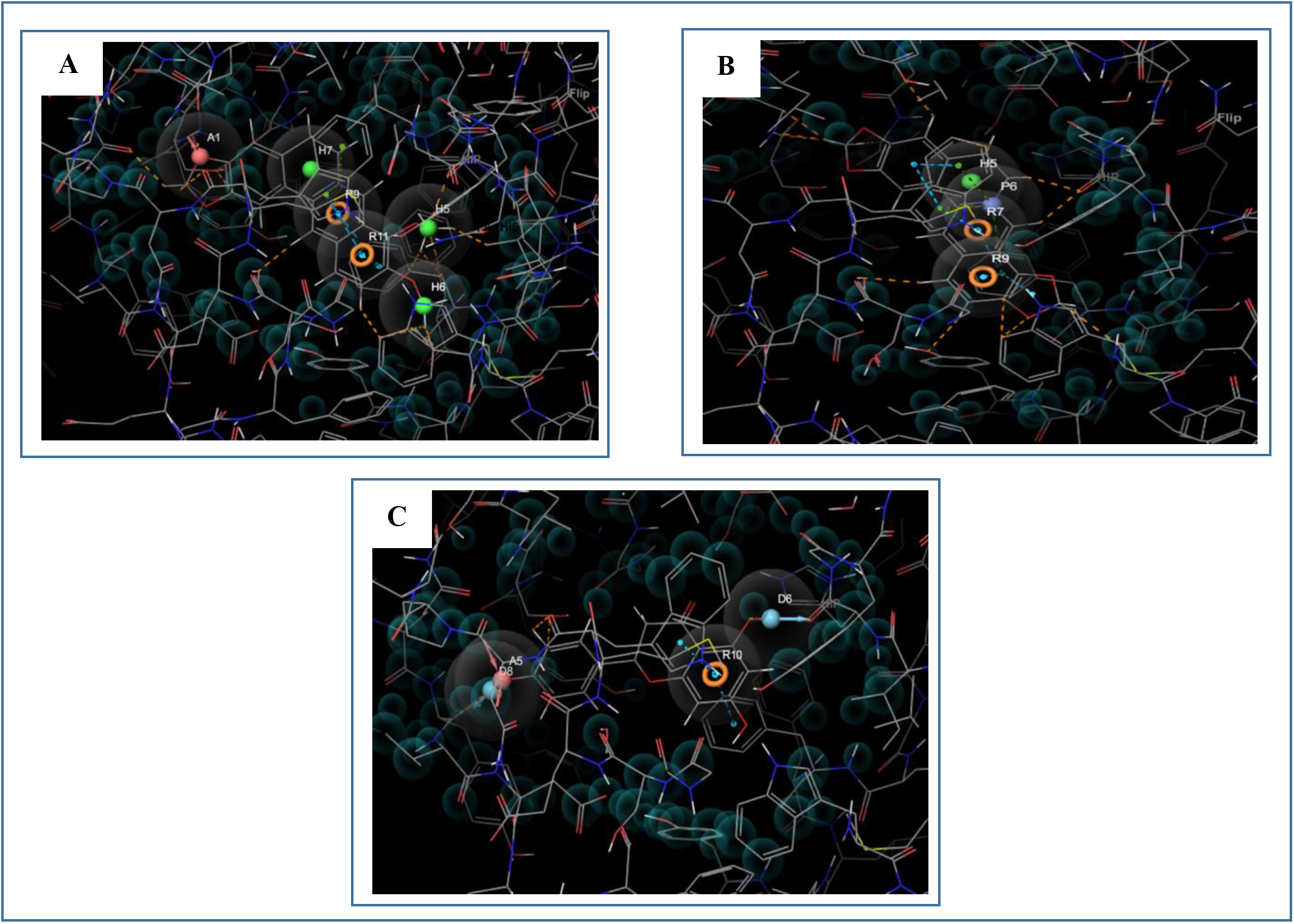
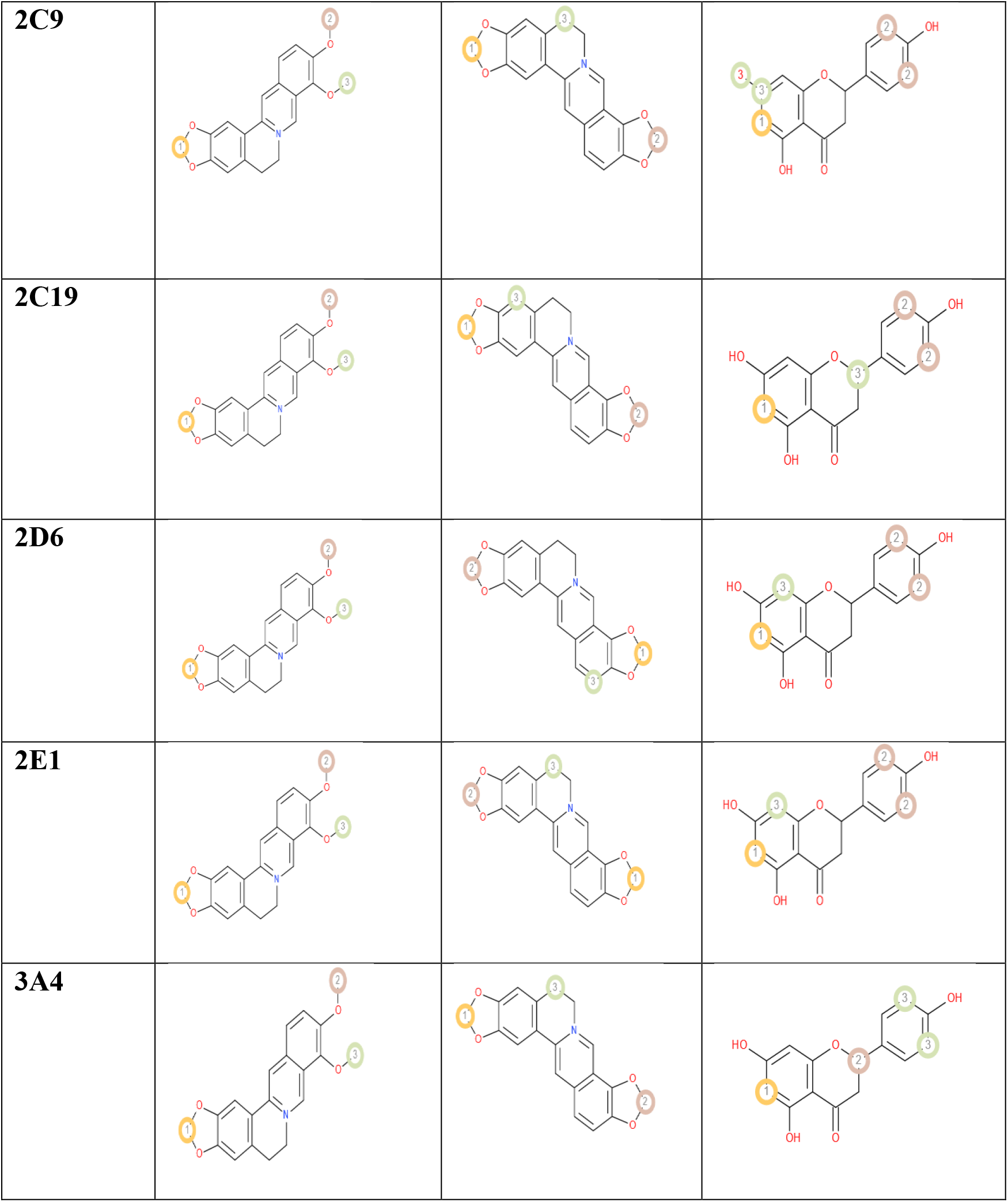
2D representation of the pharmacophore modelling of the three best ligand molecules. A. the hypothesis for berberine, B. the hypothesis for coptisine, C. the hypothesis for naringenin. The interactions between the ligands and the receptor in the hypotheses were presented by dotted dashed lines, yellow colour- hydrogen bonds, blue colour- pi-pi stacking interaction and green colour- pi-cation interaction. The bad contacts between the ligands and the pharmacophore are represented by the dotted orange lines.

**Figure 10.**
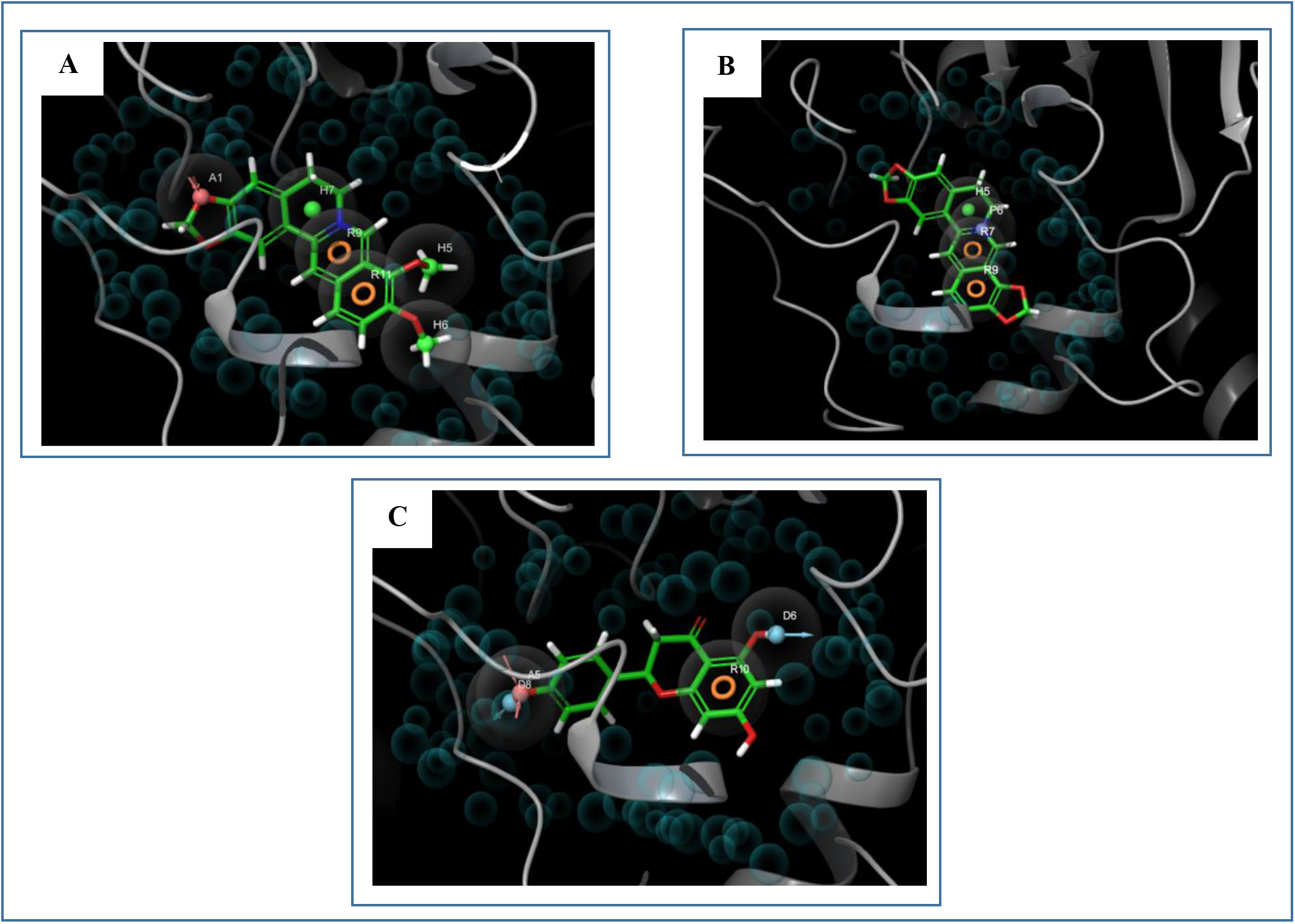
3D representation of the pharmacophore modelling of the three best ligand molecules. A. the hypothesis for berberine, B. the hypothesis for coptisine, C. the hypothesis for naringenin.

### 2.8. DFT calculation

For the Density functional theory or DFT calculation, the prepared ligands were used for DFT calculation using the Jaguar panel of Maestro Schrödinger Suite 2018-4 ^[57]^. In DFT calculation, Becke’s three-parameter exchange potential and Lee-Yang-Parr correlation functional (B3LYP) theory with 6-31G* basis set, were used ^[58, 59]^. Quantum chemical properties such as surface properties (MO, density, potential) and Multipole moments were calculated along with HOMO (Highest Occupied Molecular Orbital) and LUMO (Lowest Unoccupied Molecular Orbital) energy. Then the global frontier orbital was analyzed and hardness (**η**) and softness (**S**) of selected molecules were calculated using the following equation as per Parr and Pearson interpretation and Koopmans theorem ^[60, 61]^. The DFT calculation was done for the 3 best ligand molecules. The result of DFT calculation is summarized in **Table 10** and **Figure 11**.

**Figure 11.**
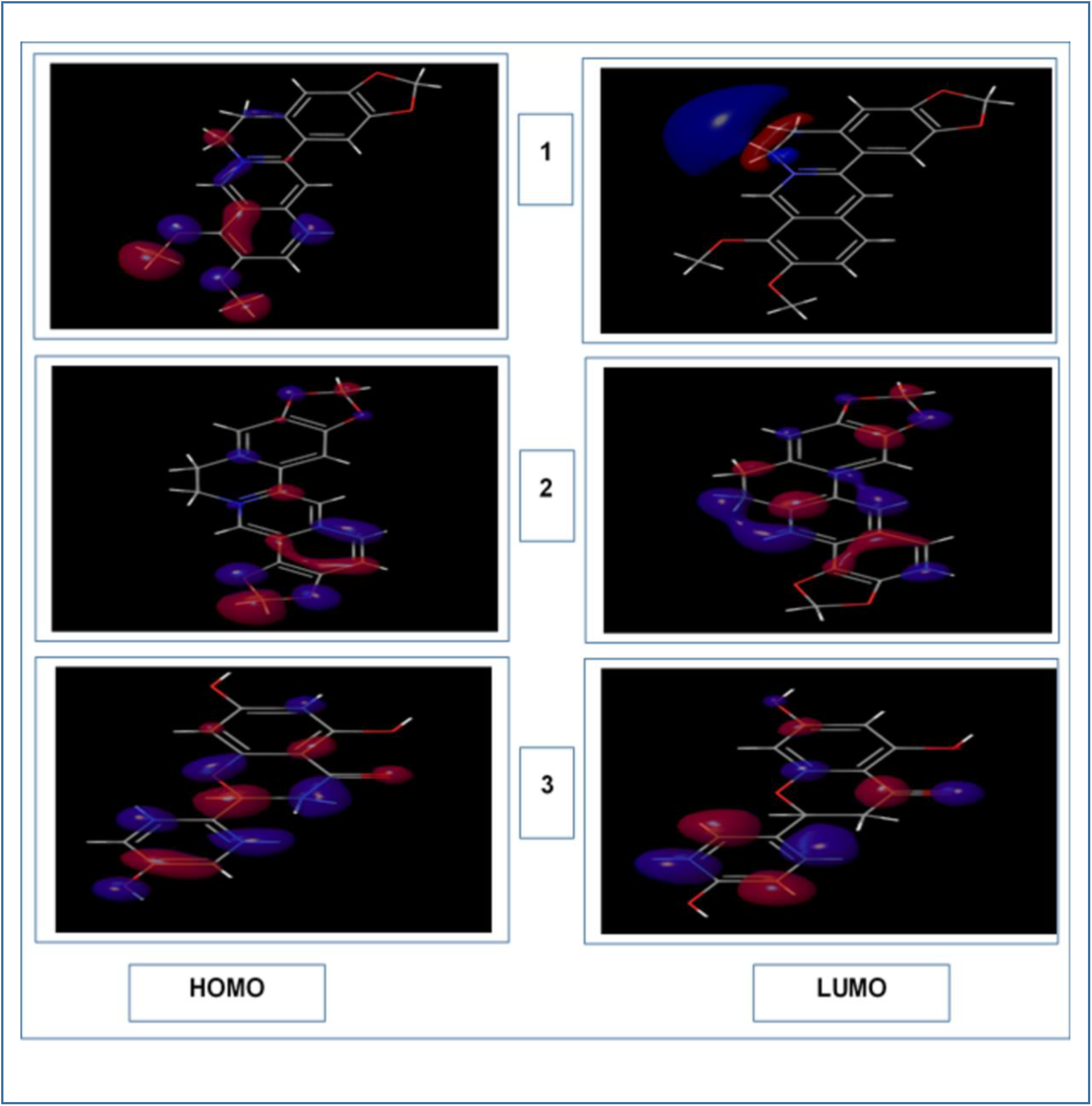
The results of DFT calculations (HOMO-LUMO structures) of, 1. Berberine, 2. Coptisine, 3. Naringenin. The HOMO structures are illustrated in the left column and the LUMO structures are illustrated in the right column.

**Figure 12.**
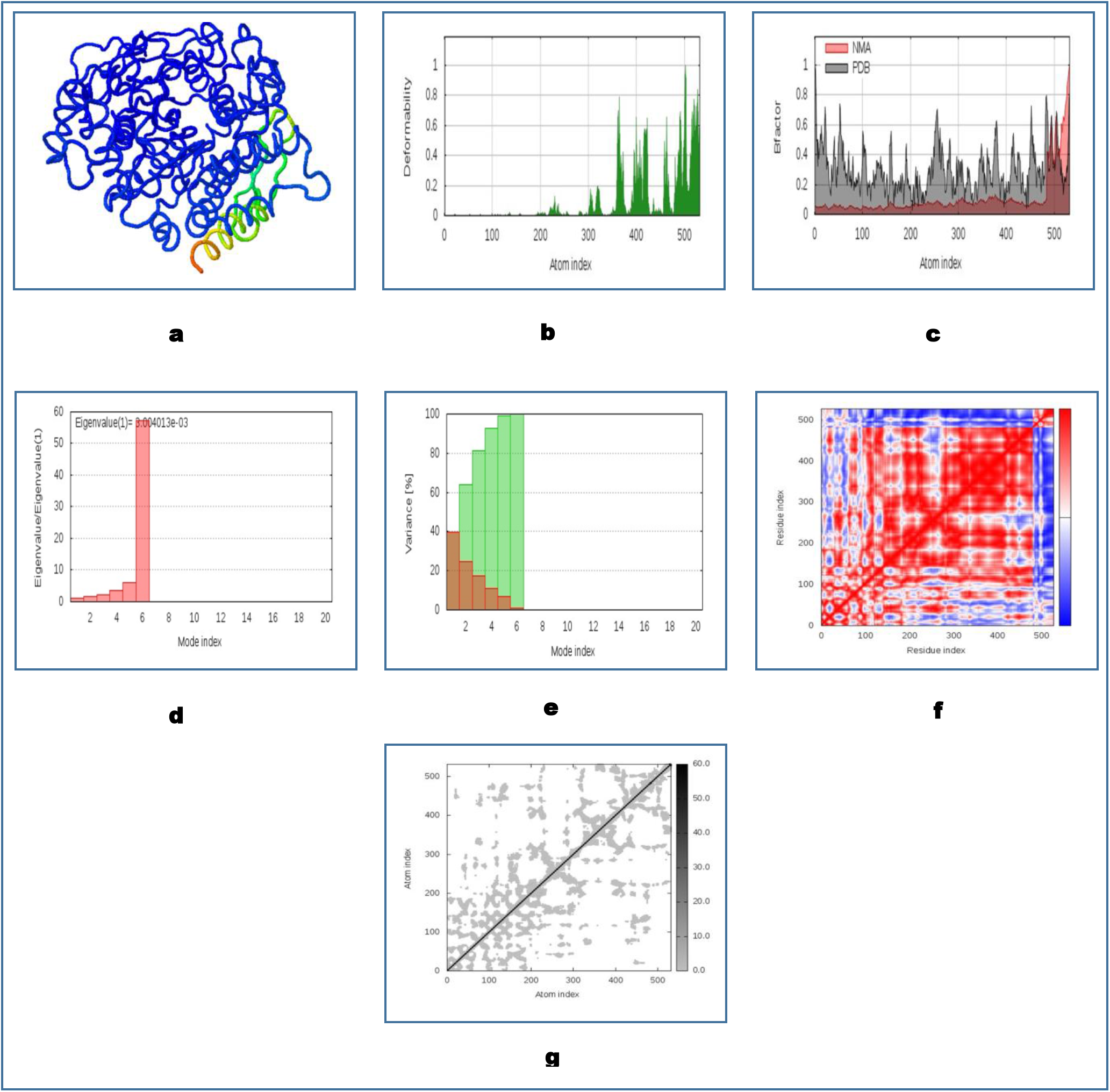
Results of molecular dynamics simulation of coptisine-AChE docked complex. (a) NMA mobility, (b) deformability, (c) B-factor, (d) eigenvalue, (e) variance (red color indicates individual variances and green color indicates cumulative variances), (f) co-variance map (correlated (red), uncorrelated (white) or anti-correlated (blue) motions) and (g) elastic network (darker grey regions indicate more stiffer regions) of the complex.

**Table 10.** The results of the DFT calculations of the best three ligands, berberine, coptisine and naringenin.

| Compound name | HOMO<br>energy (eV) | LUMO<br>energy (eV) | Gap (eV) | Hardness ($\eta$)<br>(eV) | Softness (S)<br>(eV) | Dipole<br>moment<br>(Debye) |
| --- | --- | --- | --- | --- | --- | --- |
| Berberine | -0.145 | -0.091 | 0.054 | 0.027 | 37.037 | 8.050 |
| Coptisine | -0.005 | -0.094 | 0.089 | 0.045 | 22.222 | 2.277 |
| Naringenin | -0.134 | -0.087 | 0.047 | 0.024 | 41.667 | 6.459 |

**n = (HOMOε-LUMOε)/2**,

**s = 1/ η**

### 2.9. Molecular Dynamics Simulation Study

The molecular dynamics simulation study was carried out for to determine the stability of the prepared AChE protein with the best ligand among the 14 agents. From the analysis of the results, it was declared that, coptisine was the best ligand among the selected ligand molecules. The molecular dynamics simulation study was performed by the online server iMODS (http://imods.chaconlab.org/). The server is a fast, user-friendly and effective molecular dynamics simulation tool that can be used efficiently to investigate the structural dynamics of the protein complexes. The server provides the values of deformability, B-factor (mobility profiles), eigenvalues, variance, co-variance map and elastic network. For a complex or protein, the deformability depends on the ability to deform at each of its amino acid residues. The eigenvalue has relation with the energy that is required to deform the given structure and the lower eigenvalue represents easier deformability. Moreover, the eigenvalue also represents the motion stiffness of the protein complex. The server is a fast and easy server for determining and measuring the protein flexibility ^[62, 63, 64, 65, 66]^. For analysing the molecular dynamics simulation of the three complexes, the docked PDB files were uploaded to the iMODS server and the results were displayed keeping all the parameters as default.

In this experiment, the two controls were used in molecular docking study, druglikeness property experiment and ADME/T test to compare their results with the three best selected ligands. However, the PASS prediction, P450 SOM prediction, pharmacophore mapping and modelling, solubility prediction and DFT calculations were carried out to determine and compare the biological activities of the three best ligands, for this reason, in these prediction tests, the two controls were not used.

## 3. Result

### 3.1. Molecular docking, MM-GBSA study and Induced Fit Docking

All the selected ligand molecules were docked successfully against their target, AChE. The ligand molecules that had the lowest binding energy or docking score, were considered as the best ligand molecules in inhibiting the target receptor as the lower binding energy corresponds to higher binding affinity ^[67, 68]^.

From 14 selected ligand molecules, 3 ligands were selected as the best ligands based on the lowest SP and XP docking scores or binding energies (**Table 02**). Coptisine gave the lowest SP and XP docking scores, the second lowest SP and XP docking scores were showed by berberine and the third lowest scores were given by naringenin. For this reason, these three ligands were selected as the best ligands for further analysis. In the MM-GBSA study, coptisine generated the lowest ΔG_Bind_ score of −49.91 Kcal/mol. The second lowest score was generated by berberine (−35.80 Kcal/mol) and the highest ΔG_Bind_ score was found to be −23.73 Kcal/mol, generated by naringenin. Berberine, coptisine and naringenin gave glide energies of −23.549 Kcal/mol, −16.197 Kcal/mol and −45.657 Kcal/mol, respectively. Furthermore, coptisine also generated the lowest XP G_Score_ of −14.942 Kcal/mol as well as the lowest IFD score of −1158.410 Kcal/mol. Donepezil and galantamine had ΔG_Bind_ scores of −47.06 and −5.19 Kcal/mol, respectively, XP G_Score_ of −8.209 and −5.748 Kcal/mol, respectively and IFD scores of −1148.970 and −1147.050 Kcal/mol, respectively. Berberine with XP G_Score_ of −12.889 Kcal/mol and IFD score of −1154.880 Kcal/mol, was the second lowest score generator and the highest XP G_Score_ and IFD score were showed by naringenin of −10.078 Kcal/mol and −1152.980 Kcal/mol, respectively. Berberine formed 1 carbon bond with Gln 69, 1 carbon bond and 1 conventional bond with Gly 123, 1 carbon, 1 pi-sigma, 6 pi-pi stacked and 2 pi-cation bonds with Trp 84 and 2 pi-pi stacked bonds and 1 pi-cation with Phe 330. Coptisine generated 1 pi-lone pair bond with Trp 432, 5 pi-pi stacked and 2 pi-cation bonds with Trp 84, 2 pi-pi stacked bonds and 1 pi-cation bond Phe 330, 2 carbon bonds with His 440, 1 carbon bond with Ser 122 and 1 carbon bond with Gly 117. Moreover, naringenin formed 1 conventional bond and 1 carbon bond with His 440, 1 pi-pi stacked bond with Phe 330, 1 conventional bond with Tyr 70, 1 carbon bond with Pro 86 and 2 pi-pi stacked bonds with Trp 84. The three best ligands with their respective docking score, glide energy, interacted amino acids, types of bonds and bond distances, are listed in **Table 03**.

### 3.2. Druglikeness properties

Druglikeness property experiments were conducted only for the best three ligand molecules: berberine, coptisine and naringenin (**Table 03**). Lipinski’s rule of five demonstrates that the acceptable ranges of the best drug molecule for all the five parameters are: molecular weight: ≤500, number of hydrogen bond donors: ≤5, number of hydrogen bond acceptors: ≤10, lipophilicity (expressed as LogP): ≤5 and molar refractivity from 40 to 130 ^[69]^. All the three ligands followed the Lipinski’s rule of five. Berberine, coptisine and naringenin have molecular weights less than 500 (336.36 g/mol, 320.32 g/mol and 272.25 g/mol, repectively), only naringenin had 3 hydrogen bond donors and the other two ligands didn’t have any hydrogen bond donor and both berberine and coptisine has 4 hydrogen bond acceptors, each and naringenin had 5 hydrogen bond acceptors. The logP values of berberine, coptisine and naringenin were 2.53, 2.40 and 1.84, respectively, that were also well within the accepted range of the Lipinski’s rule of five. Moreover, the molar refractivity of berberine, coptisine and naringenin were 94.87, 87.95 and 71.57, respectively. The LogS values generated by berberine, coptisine and naringenin were −4.55, −4.52 and −3.49, respectively. However, all of the ligand molecules followed the Ghose, Veber, Egan and Muegge rules and all of them showed the similar bioavailability score of 0.55. Berberine, coptisine and naringenin gave synthetic accessibility (SA) scores of 3.14, 2.96 and 3.01, respectively. Moreover, both berberine and coptisine gave topological polar surface are (TPSA) score of 40.80 Å^2^. Coptisine didn’t have any rotatable bond, berberine had 2 rotatable bonds and naringenin had 1 rotatable bond. However, the druglikeness score, drug score, solubility, reproductive effectiveness, irritant properties, tumorigenic and mutagenic properties were not available for both berberine and coptisine. Only naringenin generated results in these experiments. It gave druglikeness score of 1.9, drug score of 0.84, solubility score of −2.64 and it is not reproductive effective, irritant, tumorigenic and mutagenic. Naringenin generated quite good scores in the druglikeness property experiments. Donepezil and galantamine also showed quite good results with no violation of the Lipinski’s rule of five, Ghose, Veber, Egan and Muegge rules. They had molecular weights of 379.49 g/mol and 287.35 g/mol, respectively and druglikeness score of 7.29 and 6.2, respectively. None of them were reproductive effective, irritant, tumorigenic and mutagenic. The druglikeness properties of the three ligands and the controls are listed in **Table 04**.

### 3.3. ADME/T tests

The ADME/T tests were performed only for the three best selected ligands. All the ligands showed optimal Caco-2 permeability and all of them were p-glycoprotein non-inhibitor. However, only berberine was the p-glycoprotein substrate. Both berberine and coptisine were found to be HIA negative, which means that both of them might not be absorbed well by human intestine. All the three ligands had low plasma protein binding ability and all of them showed blood-brain-barrier crossing ability. All the three ligands were inhibitors as well as substrates for CYP450 1A2. However, only coptisine was non-inhibitor for CYP450 3A4 and both berberine and coptisine were substrates for CYP450 3A4. Moreover, all the three ligands were predicted to be non-inhibitors and non-substrates for CYP450 2C9. Although, all of them were non-inhibitors for CYP450 2C19 and only naringenin was non-substrate for CYP450 2C19. Furthermore, only naringenin was predicted to be non-inhibitor and non-substrate for CYP450 2D6. The half-life (T1/2) values of berberine, coptisine and naringenin were 1.9, 1.8 and 0.9 hours, respectively. All the three ligands showed hERG blocking capability and berberine had human hepatotoxic ability. However, all of them were not Ames mutagenic, although all of them showed the capability to do drug induced liver injury (DILI positive). Both the controls showed inhibitory activities to CYP450 2D6 as well as substrate activities to CYP450 2D6. Both of them were also substrate to CYP450 3A4. Both of them were human hepatotoxic as well as Ames negative and DILI negative. Only donepezil was hERG blocker among the two controls. The results of ADME/T tests are listed in **Table 05**.

### 3.4. PASS prediction and P450 site of metabolism (SOM) prediction

When analyzed by the admetSAR server, both berberine and coptisine were predicted to have the acute oral toxicity class of 3, for this reason, they had the same predicted LD50 value (greater than 500 mg/kg but less than 5000 mg/kg). However, naringenin had the acute oral toxicity class of 2, as a result, they had the LD50 value of greater than 50 mg/kg but less than 500 mg/kg ^[53]^.

All the three best ligands were in toxicity class 4. The prediction of activity spectra for substances (PASS prediction) study for all the three ligands were conducted to predict 10 intended biological activities and 5 intended adverse and toxic effects. To carry out the PASS prediction experiment, Pa > 0.7 was kept, since this threshold give highly reliable prediction ^[50]^. The PASS prediction results of all the three selected ligands are listed in **Table 06 and Table 07**. However, at Pa > 0.7, the intended biological activities and the adverse and toxic effects for berberine and coptisine were not generated by the PASS-Way2Drug server. Only naringenin showed the intended biological activities: membrane integrity agonist, HMOX1 expression enhancer, chlordecone reductase inhibitor, HIF1A expression inhibitor, histidine kinase inhibitor etc. Moreover, adverse and toxic effects showed by naringenin were: vascular toxicity, inflammation, hematemesis, nephrotoxicity and shivering actions.

The possible sites of metabolism by CYPs 1A2, 2A6, 2B6, 2C19, 2C8, 2C9, 2D6, 2E1 and 3A4 of berberine, coptisine and naringenin were also determined. The possible sites of a chemical compound, where the metabolism by the isoforms of CYP450 enzymes may be taken place, are indicated by circles on the chemical structure of the molecule ^[51]^. The possible sites of P450 metabolism are illustrated in **Table 08**.

### 3.5. Pharmacophore mapping and modelling

Berberine, coptisine and naringenin gave almost similar fit scores of 2.765, 2.273 and 2.637, respectively, in the pharmacophore mapping experiment. Berberine generated the normalized fit score of 0.928 and z’-score of 1.088. Coptisine showed normalized fit score of 0.757 and z’-score of 0.475. And naringenin had the normalized fit score of 0.659 and z’-score of 0.749. Furthermore, hydrophobic centres generated by berberine, coptisine and naringenin were 1, 1 and 3 respectively. Both berberine and coptisine had 2 hydrogen bond acceptors each, whereas, naringenin didn’t generate any hydrogen bond acceptor. Moreover, only naringenin had 1 hydrogen bond donor. However, none of the molecules showed positively charged centre, negatively charged centre and aromatic ring (**Figure 08 and Table 09**).

The three best ligands were used to generate pharmacophore hypotheses. Berberine generated 6 point hypothesis and both coptisine and naringenin showed 4 point hypothesis each. Berberine generated 2 pi-cation bonds, 4 pi-pi stacked interactions and 2 hydrogen bonds with its pharmacophore. Although it generated several good contacts (not shown here), however, it also generated 13 bad contacts with its pharmacophore. Coptisine showed 2 hydrogen bonds, 6 pi-pi stacked interactions and 2 pi-cation bonds. Like berberine, coptisine also generated several good contacts, however, it also showed 12 bad contacts with its pharmacophore. Naringenin showed 2 pi-pi stacked interactions, 2 hydrogen bonds and 2 bad contacts with the pharmacophore. Moreover, it also generated a good number of good contacts with the pharmacophore of AChE. None of the ligands generated ugly contact with AChE (**Figure 09** and **Figure 10**).

### 3.6. DFT Calculation

In the DFT calculations, berberine showed HOMO energy of −0.145 eV, LUMO energy of −0.091 eV and gap energy of 0.054 eV as well as the dipole moment of 8.050 debye. Coptisine generated HOMO and LUMO energies of −0.005 eV and −0.094 eV, respectively and gap energy of 0.089 eV. On the other hand, naringenin gave HOMO energy of −0.134 eV, LUMO energy of −0.087 eV, gap energy of 0.047 eV and the dipole moment of 6.459 debye (**Table 10** and **Figure 11**).

### 3.7. Molecular Dynamics Simulation

**Fig. 12a** illustrates the normal mode analysis (NMA) of the prepared enzyme-ligand complex. The deformability graphs of the three complexes illustrate the peaks in the graphs correspond to the regions in the protein with deformability (**Fig. 12b**). The B-factor graph of the complex gives easy visualisation and understanding of the comparison between the NMA and the PDB field of the complexes (**Fig. 12c)**. The eigenvalue of the complex is illustrated in **Fig**. **12d**. The docked complex with the prepared protein generated eigenvalue of 3.004013e-04. The variance graph indicates the individual variance by red coloured bars and cumulative variance by green coloured bars (**Fig. 12e**). **Fig. 12f** illustrate the co-variance map of the complexes where the correlated motion between a pair of residues are indicated by red colour, uncorrelated motion is indicated by white colour and anti-correlated motion is indicated by blue colour. The elastic map of the complex shows the connection between the atoms and darker grey regions indicate stiffer regions (**Fig. 12g**) ^[63, 64, 65]^.

## 4. Discussion

The aim of docking experiment is to determine the best possible pose of a ligand molecule within the constraint of binding pocket of a receptor and thereafter, a binding energy is calculated. The lower the binding energy (docking score), the higher the affinity of binding and vice versa. In the experiment, total 14 ligand molecules were selected to act against the AChE enzyme which is responsible for AD development. Each of the 14 ligands was docked against the target receptor to evaluate their anti-AChE activity and from the experiment, three best ligands were selected for further analysis. The best possible ligand molecules were selected based on their binding energy, where the lower bind energy was preferred. Coptisine gave the lowest SP docking score of −10.148 Kcal/mol, so coptisine should be the best molecule to inhibit AChE. Furthermore, coptisine also generated the lowest XP docking score or binding energy of −15.560 Kcal/mol. Moreover, the second lowest and the third lowest SP and XP docking scores were given by berberine (−9.658 Kcal/mol and −13.571 Kcal/mol, respectively) and naringenin (−9.266 Kcal/mol and −9.342 Kcal/mol, respectively), respectively. For this reason, these three molecules were selected as the best three ligand molecules among the 14 ligands. The MM-GBSA study was carried out for only the three best ligands that showed the best results in the docking study. In the MM-GBSA study, the most negative ΔG_Bind_ score (the lowest score) is considered as the best ΔG_Bind_ score ^[70]^. IFD study is performed to understand the accurate binding mode and to ensure the accuracy of active site geometry. XP G_Score_ is generated in the IFD experiment, which is an empirical scoring function that estimates the ligand binding free energy. The lowest value of XP G_Score_ is considered as the best value and is always appreciable ^[71, 72, 73, 74]^. In the MM-GBSA study, based on the ΔG_Bind_ score, XP G_Score_ value and IFD score, coptisine can be considered as the best ligand molecule among the selected three ligands. Further analysis showed that berberine interacted with 4 amino acids (Gln 69, Phe 330, Gly 123 and Trp 84), whereas, coptisine interacted with 6 amino acids (Trp 432, Trp 84, Phe 330, His 440, Ser 122 and Gly 117) and naringenin interacted with 5 amino acids (His 440, Phe 330, Tyr 70, Pro 86 and Trp 84). When the three best ligands were compared with the positive controls, it was observed that the performances of both donepezil and galantamine in the docking studies were less satisfactory than berberine, coptisine and naringenin. For this reason, it can be concluded that the three best selected ligand molecules showed superior performances in the molecular docking studies. The list of the amino acids take part in the interaction of the ligands and the positive controls with AChE are listed in **Table 02**. Trp 84 and Phe 330 amino acids were found to be the most commonly interacting amino acids of AchE with all the 14 ligands and the positive control galantamine. Moreover, His 440 and Gly 117 were also found to be quite commonly interacting amino acids with most of the selected ligands. Again, only naringenin was found to have interacted with Tyr 70 and carvacrol was found in interaction with Arg 289, other than another positive control, donepezil. Some amino acids were found to be common in interaction with some ligands but they were not found in the controls, for example: Ile 439, which interacted with both estragole and harmaline, but didn’t form any interaction with the positive controls. Again, Trp 279 of AChE was only found to be interacted with the positive control donepezil which was not found in interaction with other ligands. The Ile 444 (interacted with myrtenal) and Tyr 130 (interacted with scoulerine) were very unique amino acids which were not found to be interacted with any other ligands and positive controls. However, other interacting amino acids of galantamine, Glu 199, Tyr 121 and Tyr 70 were also found to be in interaction with some of the ligands. AChE possesses several notable amino acids in its active-site gorge i.e., Ser 200, Glu 327, His 440, Trp 279, Tyr 121, Phe 330 and Trp 84. All the 14 ligands were predicted to interact with both Phe 330 and Trp 84 and most of them also interacted with His 440 by hydrogen and hydrophobic interactions, it can be concluded that all of the 14 ligands were found to bind with the AChE receptor at its active site. The hydrogen and hydrophobic interactions are important for strengthening the receptor-ligand interactions ^[75, 76]^.

Estimating the drug likeness properties facilitates the drug discovery and development processes. The molecular weight and topological polar surface area (TPSA) influence the drug permeability through the biological barrier. The higher the molecular weight and TPSA values, the lower the permeability of the drug molecule is and vice versa. Lipophilicity is expressed as the logarithm of partition coefficient of a drug molecule in organic and aqueous phase (LogP). It affects the absorption of the drug molecules in the body and higher LogP corresponds to lower absorption and vice versa. LogS value influences the solubility of a drug molecule and the lowest value is always appreciable. The number of hydrogen bond donors and acceptors above the acceptable ranges also affects the capability of a drug molecule to cross the cell membrane. The number of rotatable bonds affects the druglikeness properties and the acceptable range is <10. Moreover, the Lipinski’s rule of five demonstrates that a successful drug molecule should have properties within the acceptable range of the five Lipinski’s rules ^[68, 77, 78]^. The druglikeness property experiment was conducted for the three best ligand molecules. Moreover, according to the Ghose filter, a candidate drug molecule should have LogP value of −0.4 to 5.6, molecular weight between 160 and 480, the total number of atoms from 20 to 70, molar refractivity from 40 to 130, to qualify as a successful drug ^[79]^. Veber rule describes that the oral bioavailability of a possible drug molecule depends on two factors i.e., the polar surface are which should be equal to or less than 140 Å^2^ and 10 or fewer numbers of rotatable bonds ^[80]^. Furthermore, according to the Egan rule, the absorption of a candidate drug molecule also depends on two factors: the polar surface area (PSA) and AlogP98 (the logarithm of partition co-efficient between n-octanol and water) ^[81]^. And according to the Muegge rule, for a drug like chemical compound to become a successful drug molecule, it has to pass a pharmacophore point filter, which was developed by the scientists ^[82]^. Moreover, how easily a target compound can be synthesized is determined by the synthetic accessibility (SA) score. The score 1 represents very easy to synthesize, whereas, the score 10 represents very hard to synthesize ^[83]^. The bioavailability score describes the permeability and bioavailability properties of a possible drug molecule ^[84]^.

Berberine, coptisine and naringenin had molecular weights of 336.36 g/mol, 320.32 g/mol and 272.25 g/mol, respectively. As the lower molecular weight is always appreciable, naringenin should be the best among the three ligands. Both berberine and coptisine showed the TPSA value of 40.80 and naringenin had TPSA of 71.57. Since, the lower TPSA value always gives the good results, both berberine and coptisine performed better than naringenin. In the case of lipophilicity (expressed as LogP), the lower value is always required. Naringenin, with the lowest logP value among the three ligands (1.84), showed quite satisfactory performance in the lipophilicity experiment. The other two ligands, berberine and coptisine, with their logP values of 2.53 and 2.40, respectively, also showed quite good performances in the study. Berberine and coptisine showed almost similar LogS values of −4.55 and −4.52, respectively. The number of rotatable bonds showed by berberine (2), coptisine (0) and naringenin (1) were well within the acceptable range. All the best three ligands were predicted to follow the Lipinski’s rule of five.

The three ligands also found to follow the Ghose filter, Veber, Egan and Muegge rules. Considering all the aspects of druglikeness property experiment, it can be concluded that, all the three best ligand molecules performed quite similarly in the druglikeness property experiment. When compared with the controls, it was observed that all the three ligands showed quite sound performances in the druglikeness property experiment.

ADME/T tests are carried out to determine the pharmacological and pharmacodynamic properties of a candidate drug within a biological system. For this reason, it is a crucial determinant of the success of a drug research and development. Blood brain barrier (BBB) is the most important factor for the drugs that primarily target the brain cells. Moreover, since most of the drugs are administered through the oral route, it is required that these drugs should be absorbed well in the intestinal tissue. p-glycoprotein (p-gp) in the cell membrane aids in transporting many drugs inside the cell. Therefore, the inhibition of p-gp affects the drug transport. Caco-2 cell line is widely used in *in vitro* study of drug permeability tests. Its permeability decides that whether the drug will be easily absorbed in the intestine or not. Orally absorbed drugs travel through the blood circulation and deposit back to liver. In the liver, the enzymes of Cytochrome P450 family metabolize the drugs and excrete the metabolized drugs through bile or urine. As a result, inhibition of any one of these enzymes affects the biodegradation of the drug molecule ^[85, 86]^. Moreover, if a compound is found to be substrate for one or more isoforms of CYP450 enzymes, then that compound is predicted to be metabolized well by the respective CYP450 enzyme or enzymes ^[87]^. The binding of drugs to plasma proteins is also a crucial pharmacological parameter that affects the pharmacodynamics, excretion and circulation of drugs. A drug’s proficiency is depended on the degree of its binding with the plasma protein. A drug can cross the cell layers or diffuse easily if it binds to the plasma proteins less efficiently and vice versa ^[88]^. Human intestinal absorption (HIA) is a very important process for the orally administered drugs. It depicts the absorption of orally administered drugs from the intestine into the bloodstream ^[89, 90, 91]^. Drug half-life describes the time it takes for the amount of a drug in the body to be reduced by half or 50%. The greater the half-life of a drug, the longer the drug would stay in the body and the greater its potentiality. For this reason, half-life determines the doses of drugs ^[92, 93, 94]^. HERG is a K+ channel found in the heart muscle which mediates the correct rhythm of the heart. HERG can be blocked by certain drugs which may lead to the cardiac arrhythmia and death ^[95, 96]^. Being the main site of metabolism, human liver is extremely vulnerable to the harmful effects of various drugs and xenobiotic agents. Human hepatotoxicity (H-HT) indicates any type of injury to the liver that may lead to organ failure and even death. Human hepatotoxicity, sometimes, is also responsible for the withdrawal of approved drugs from the market ^[97, 98]^. Ames test is a mutagenicity test that is used to detect the potential mutagenic chemicals which have the capability to cause mutations and cancer ^[99]^. Drug induced liver injury (DILI) is the injury to the liver that are caused by administration of drugs ^[100]^. The results of ADME/T test are listed in **Table 05**.

In the absorption section, all the three ligands performed quite similarly. All the ligands showed optimal Caco-2 permeability and all of them were non-inhibitors of p-gp. For this reason, none of them inhibited the actions carried out by p-gp. However, only berberine was the p-gp substarte, as a result, berberine should be taken up by the cell more easily than the other two ligands. Again, since only naringenin showed HIA capability, it should be absorbed well by the human intestine. In the absorption section, all the ligands showed quite similar and sound performances. In the distribution section, berberine and coptisine showed low plasma protein binding capability than naringenin. However, all of them were capable of crossing the blood brain barrier. In the metabolism section, naringenin showed the poorest performances. Berberine was the inhibitor of CYP450 1A2, CYP450 3A4 and CYP450 2D6. Moreover, the ligand was also a substrate for CYP450 1A2, CYP450 3A4, CYP450 2C19 and CYP450 2D6. As berberine was found to be the substrate of these enzymes, they might metabolize the ligand very efficiently. Coptisine was inhibitor for CYP450 1A2 and CYP450 2D6. However, since it was predicted to be the substrate of CYP450 1A2, CYP450 3A4, CYP450 2C19 and CYP450 2D6 enzymes like berberine, these enzymes may also metabolize coptisine effectively. Naringenin was the inhibitor of CYP450 1A2 and CYP450 3A4. On the other hand, it was the substrate for only CYP450 1A2, for this reason, naringenin was predicted to be metabolized only by CYP450 1A2. In the metabolism section, berberine and coptisine showed quite good as well as almost similar results. However, naringenin showed unsatisfactory performance in the metabolism section. In the excretion section, naringenin showed the lowest half-life of 0.9 hours. Therefore, it can be declared that, naringenin’s performances were not so satisfactory in the excretion section. In the toxicity section, all the three ligands were hERG blockers, however, all of them proved to be safe in the Ames mutagenicity test. On the other hand, since all of them were DILI positive, they could cause liver injuries. Moreover, coptisine and naringenin were not human hepatotoxic, whereas, berberine was a hepatotoxic agent. Their performances were quite good when compared with the two controls used in the experiment. Furthermore, coptisine showed even better performances than the positive controls in some aspects of the experiment. Comparing with the two controls, it can be declared that, the three best ligands performed well in the docking study, druglikeness property experiment and ADME/T test. All the three best ligands showed satisfactory results in these experiments. Moreover, coptisine generated even superior results than the controls in some aspects of the experiment.

Prediction of Activity Spectra for Substances or PASS prediction is carried out to estimate the possible biological activities associated with drug-like molecules. The PASS method estimates the probabilities based on the structures of the compounds and their molecular mass. Two parameters are used for the PASS prediction: Pa and Pi. The Pa is the probability of a compound “to be active” and Pi is the probability of a compound “to be inactive”. The values of both Pa and Pi can range from zero to one ^[49]^. If the value of Pa is greater than 0.7, the probability of exhibiting the activity of a substance in an experiment is higher. On the other hand, if the Pa is greater than 0.5 but less than 0.7, the probability of exhibiting a particular activity in an experiment is good, although less than the probability of the activity determined when Pa > 0.7 threshold is used. Moreover, if Pa is less than 0.5, the probability of exhibiting the activity is the least ^[101]^. However, the chance of finding any given activity in an experiment increases with the increasing value of Pa as well as decreasing value of Pi ^[49]^. The PASS prediction was carried out to determine 10 biological activities and 5 adverse and toxic effects of the three selected ligands. Since the intended activities were not generated by PASS-Way2Drug server (http://www.pharmaexpert.ru/passonline/) at Pa > 0.7, the PASS prediction were not demonstrated for berberine and coptisine. However, naringenin generated predictions for all the 10 biological as well as toxic activities.

ProTox-II server measures the toxicity of a chemical compound and classifies the compound into a toxicity class ranging from 1 to 6. The server classifies the compound according to the Globally Harmonized System of Classification and Labelling of Chemicals (GHS) ^[53]^. According to the GHS system, Class 1: fatal if swallowed (LD50 ≤ 5), class 2: fatal if swallowed (5 < LD50 ≤ 50), class 3: toxic if swallowed (50 < LD50 ≤ 300), class 4: harmful if swallowed (300 < LD50 ≤ 2000), class 5: may be harmful if swallowed (2000 < LD50 ≤ 5000) ^[102]^. However, ProTox-II server adds one more class to the 5 classes, making them 6 classes in total, class VI: non-toxic (LD50 > 5000) (http://tox.charite.de/protox_II/index.php?site=home) ^[53]^. All the selected three ligands were in toxicity class 4, meaning that, they might be harmful, if swallowed. The possible sites where the metabolism on a chemical structure may be carried out by the isoforms of CYP450 enzymes, are indicated by circles on the chemical structure of the molecule ^[51]^. In the P450 SOM experiment, naringenin gave the best result since it generated 4 SOMs for all the CYP450 enzymes, except CYP450 2C9 (5 SOMs). However, berberine and coptisine gave the similar results because they showed 3 SOMs for all of the CYP450 enzymes.

The pharmacophore mapping study of the three best ligand molecules were carried out by the PharmMapper server. PharmMapper generates three types of scores i.e., fit score, normalized fit score and z’-score. The target proteins (receptors) with the highest fit scores and highest normalized fit scores reflect that they should be the potential targets for a query compound to bind. Moreover, z’-score is generated form the fit score and high z’-score corresponds to high significance of the target to a query compound and vice versa ^[103, 104, 105, 106]^.

The pharmacophore mapping experiment of berberine and naringenin gave almost similar fit scores of 2.765 and 2.637, respectively. However, berberine showed the highest normalized fit score of 0.928. Coptisine and naringenin had the normalized fit scores of 0.757 and 0.659. As a result, with the highest fit score and normalized fit score, the target protein AChE should be the most potential target for berberine, among the three ligands. Since berberine also generated the highest z’-score of 1.088, the binding between berberine and AChE is the most significant among the three ligands. However, with the lowest z’-score of coptisine (0.475), the binding between coptisine and AChE is less significant than the other two ligands. None of the ligands showed any positively charged centre, negatively charged centre and aromatic ring. In the pharmacophore modelling experiment, berberine showed the best result, however, the other two ligands also showed good results in the study (**Figure 08 and Table 09**).

The Phase pharmacophore perception engine is a tool of Maestro-Schrödinger Suite 2018-4 which is used for pharmacophore modelling, QSAR model development and screening of 3D database. The engine provides 6 types of built-in features and the pharmacophore modelling is mainly done based on these 6 types of features i.e., hydrogen bond acceptor (A), hydrogen bond donor (D), negative ionizable (N), positive ionizable (P), hydrophobe (H), and aromatic ring (R). However, the number of features can be increased by customization. The pharmacophore modelling generates a hypothesis which can be used successfully in biological screening for further experiments ^[107]^.

All the three best ligand molecules successfully generated the pharmacophore modelling hypothesis with AChE. In generating the hypothesis, 6 features were selected for berberine, 4 features were selected for coptisine and 4 features were selected for naringenin. For this reason, berberine showed 6 point hypothesis and both coptisine and naringenin generated 4 point hypothesis. All the three best ligands showed quite sound results in their pharmacophore modelling hypotheses, as a result, all of these hypotheses can be used in screening effectively (**Figure 09 and Fig. 10**).

Frontier orbitals study or DFT calculation is one of the essential methods determining the pharmacological properties of various small molecules ^[108]^. HOMO and LUMO help to study and understand the chemical reactivity and kinetic stability of small molecules. The term ‘HOMO’ describes the regions on a small molecule which donate electrons during a complex formation and the term ‘LUMO’ indicates the regions on a small molecule that receive electrons from the electron donor(s). The difference in HOMO and LUMO energy is called gap energy and gap energy corresponds to the electronic excitation energy. The compound that has the greater orbital gap energy, tends to be energetically more unfavourable to undergo a chemical reaction and vice versa ^[58, 109, 110]^. Moreover, gap energy also has correlation with the hardness and softness properties of a molecule ^[111]^. The DFT calculations were carried out for all the three best ligand molecules. Naringenin showed the lowest gap score of 0.047 eV, therefore, naringenin is energetically more favourable to undergo chemical reactions than the other two ligands. Moreover, the lowest gap scores also corresponds to lowest hardness score and the highest softness score of naringenin, 0.024 eV and 41.667 eV, respectively. However, berberine generated the highest dipole moment score of 8.050 debye (**Table 10** and **Figure 11**).

Taking all the aspects into account, all the three best ligand molecules showed almost similar results in all the experiments, except the PASS prediction experiment. Due to the unavailability of data, the PASS prediction and solubility tests for berberine and coptisine could not be determined. However, naringenin showed quite good results in PASS prediction and solubility experiments. Comparing with the two positive controls, it can be concluded that, the best three ligands performed very well in the experiments. Coptisine could be regarded as the best ligand molecule among the three selected ligands based on the docking studies (molecular docking, MM-GBSA and IFD studies) and many other aspects of the conducted experiment, although berberine and naringenin also showed quite satisfactory results. In some fields, coptisine generated far better results than the positive controls. From the molecular dynamics study of the prepared AChE- coptisine docked complex, it was clear that the prepared enzyme-ligand complex had very low chance of deformability (**Fig. 12b**) as well as it had quite high eigenvalue of 3.004013e-03, so the deformability would be quite difficult for the complex (**Fig. 12d**). However, the variance map showed high degree of cumulative variances than individual variances (**Fig. 12e**). The co-variance and elastic network map also produced quite satisfactory results (**Fig. 12f** and **Fig. 12g**). The three selected ligand molecules can be used as potential agents to treat AD.

Overall, in our study, coptisine emerged as the most potent anti-AChE agent. However, more *in vitro* and *in vivo* researches should be performed on the three best ligands to finally confirm the findings of this study.

## 5. Conclusion

14 compounds that have AChE inhibiting activities, were selected for our study to determine the three best agents among them and their potential effects, safety and efficacy by conducting various experiments i.e., molecular docking, druglikeness property experiments, ADME/T tests, PASS prediction studies and P450 site of metabolism prediction. From the experiment, three ligands i.e., berberine, coptisine and naringenin were determined to be the best three ligands to inhibit AchE. All the three ligands showed quite similar and sound results in all the aspects of the experiment. Berberine can be found in *Berberis vulgaris*, coptisine can be extracted from plants like *Coptis chinensis, Berberis bealei and Phellodendron chinense and* naringenin can found in *Citrus junos*. Since these plants contain anti-AChE agents, these plants can be used to treat the Alzheimer’s disease, naturally, by targeting the AChE pathway. Finally, this study recommends the berberine, coptisine and naringenin as the most potential anti-AChE agents among the 14 selected ligands to treat AD. Hopefully, this study will raise interest among the researchers.

## Data Availability

all the data were obtained by reviewing the literature available in google scholar

## 6. Acknowledgements

Authors are thankful to Swift Integrity Computational Lab, Dhaka, Bangladesh, a virtual platform of young researchers, for providing the tools.

## 7. Compliance with Ethical Standards

### 7.1. Conflict of interest

Bishajit Sarkar declares that he has no conflict of interest. Md. Asad Ullah declares that he/she has no conflict of interest. Md. Nazmul Islam Prottoy declares that he/she has no conflict of interest.

### 7.2. Ethical approval

This article does not contain any studies with human participants or animals performed by any of the authors.

## Notes

### Competing Interest Statement

The authors have declared no competing interest.

### Funding Statement

No funding was received for conducting the experiment.

